# The effect of a post-learning nap on motor memory consolidation in people with Parkinson’s disease: a randomised controlled trial

**DOI:** 10.1101/2025.01.09.25320277

**Authors:** Letizia Micca, Genevieve Albouy, Bradley R King, Nicholas D’Cruz, Alice Nieuwboer, Wim Vandenberghe, Pascal Borzée, Bertien Buyse, Dries Testelmans, Judith Nicolas, Moran Gilat

## Abstract

**Study Objectives:** Motor memory consolidation is a process by which newly acquired skills become stable over time in the absence of practice. Sleep facilitates consolidation, yet it remains unknown whether sleep-dependent consolidation is intact in people with Parkinson’s disease (PD). Here, we investigated whether a post-learning nap - as compared to wakefulness - improves motor memory consolidation in PD.

**Methods:** Thirty-two people with PD and 32 healthy older adults (HOA) learnt a finger-tapping sequence task before being randomised to a nap or wake intervention. Consolidation was measured as the change in performance between pre- and post-intervention and at 24-hour retention. Automaticity was measured with dual-task cost, assessed at post-intervention and at post-night. Sleep architecture and electrophysiological markers of plasticity were extracted from the post-learning nap to assess their correlation with performance changes.

**Results:** The behavioural results provided weak evidence for similar consolidation effects after sleep and wakefulness in both PD and HOA, and showed no intervention effects. Napping also did not affect dual-task costs in PD or HOA. Results suggested positive correlations between performance improvements and slow wave amplitude and slope in PD, and opposite associations between cross-frequency coupling and performance change in PD and HOA.

**Conclusions:** Findings suggest that napping did not have a beneficial effect on the consolidation of a finger-tapping task over wakefulness in either PD or HOA groups. In PD, sleep markers of plasticity were associated with performance improvements, implying that equivalent memory consolidation between HOA and PD may be differently related to sleep-related processes.

**Statement of significance:** This is the first study to directly compare the effect of post-learning napping with wakefulness on motor memory consolidation in people with Parkinson’s disease (PD), and to show no beneficial effect of sleep as compared to wakefulness. Further, motor memory consolidation may be unaffected in early-to-mid stage PD as compared to healthy older adults (HOA). In PD, electrophysiological markers of plasticity during sleep were positively related to motor performance improvement, indicating that HOA and PD may achieve equivalent performance outcomes through different consolidation mechanisms.

## Introduction

Parkinson’s disease (PD) is the second most common neurodegenerative disorder worldwide, characterised primarily by the progressive degeneration of nigrostriatal dopaminergic neurons [1]. Eventually, the striatal dopamine depletion results in the cardinal motor symptoms of bradykinesia, resting tremor and/or muscle rigidity [2]. The progressive striatal dysfunction in PD likely affects multiple aspects of motor memory, including learning during practice, consolidation, retention and automaticity [3]. Consolidation is a process that transforms recently acquired memory traces into robust and long-lasting memories in the absence of further practice [4]. Practice and consolidation are both needed to achieve retention and automaticity. Retention indicates that the learned skill is maintained (retained) and has become robust against interposed activities and time, with performance being re-assessed across various timescales (ranging from minutes to hours, days or months after learning). Automaticity refers to the ability to perform the motor task without focused attention, typically assessed with a dual-task [5]. In PD, initial motor learning is relatively spared [6], but retention and ultimately automaticity are impaired [7–9]. Reduced retention and automaticity have been demonstrated already early in PD under certain task conditions [10]. Due to these deficits, many neurorehabilitation strategies that depend on the acquisition or re-learning of motor skills and their retention (e.g., writing training or strategies for fall prevention) require continued practice and attentional resources to maintain sufficient performance levels in PD [9,11].

In healthy young adults, non-rapid eye movement (NREM) sleep, particularly stages 2 (N2) and 3 (N3) [4], is known to enhance memory consolidation of tasks engaging the hippocampo-cortical network, such as motor sequence learning (MSL) tasks [12–15]. In healthy older adults (HOA) sleep-related motor memory consolidation is diminished [16–19], but can appear under certain conditions [20–22]. In people with PD, only three prior studies measured motor performance after a period of sleep, all using MSL tasks. Interestingly, these three studies showed that sleep following practice had similar effects on consolidation as in HOA [23–25]. However, none of these studies directly compared sleep with an equivalent period of wakefulness to determine whether these effects were specifically sleep-related.

The deficit in sleep-related motor memory consolidation in HOA is likely due to age-related alterations of sleep macro-architecture, such as increased sleep fragmentation with larger percentages of wakefulness and percentage of stage 1 (N1) sleep, and reduced periods of N3 sleep [26]. The electrophysiological markers of plasticity during sleep, such as spindles, slow wave and their coupling, are also altered in HOA, with decreased amplitude of spindles and slow waves [13,16,17,27,28], and a desynchronised coupling between the two [29].

Notably, people with PD experience even more severe sleep disruptions than HOA [30]. Indeed, sleep disorders in PD, such as REM sleep behaviour disorder (RBD), disturbances with sleep onset and maintenance insomnia, as well as, excessive daytime somnolence [31–33], are prevalent and known to affect quality of life [31] and cognition [34]. Yet, the specific impact of sleep disorders on motor memory consolidation in PD remains unclear. While sleep spindles and slow waves exhibit reduced amplitudes in PD compared to HOA [34], a recent case-control study surprisingly found stronger coupling between spindles and slow waves in PD compared to HOA [35]. Additionally, greater spindle density was associated to better overnight consolidation of a verbal pair-association cognitive task in people with PD [36].

Taken together, sleep appears to play a role in offline consolidation processes in PD, but whether it is impaired – relative to HOA – remains an open question. Specifically, direct evidence comparing the effects of sleep and wakefulness on motor memory consolidation is lacking. Hence, with this study we primarily aimed to determine whether a 2-hour post-learning nap would improve the consolidation of a MSL finger-tapping task, as compared to a similar period of post-learning wakefulness, in PD and HOA. Moreover, we set out to study the effect of post-learning napping on the retention of motor performance at a 24-hour retest, and on task automaticity measured with a cognitive dual-task. We expected that napping would result in a similar offline performance changes immediately after the intervention, at 24-hours retest, and in better dual-task costs, as compared to wakefulness in both PD and HOA. We further anticipated that while people with PD would show reduced performance after 24 hours compared to HOA, a post-learning period of sleep may contribute to maintaining performance gains on the MSL task. We also explored the relationship between performance changes and electrophysiological sleep features for the participants in the nap group, to identify potential mechanisms occurring during sleep, which could explain the expected similarity in sleep-related consolidation.

## Methods

This phase-II study utilised a randomised controlled open-label design (PD: N= 32, HOA: N = 32). The procedures and hypotheses were pre-registered on clinicaltrials.gov (Experiment 1, clinicaltrials.gov/study/NCT04144283), and the reporting adheres to the Consolidated Standards of Reporting Trials (CONSORT) guidelines (see Appendix A) [37]. An independent researcher performed randomisation to either the nap or wake intervention group using a computerised blocked randomisation sequence (app.studyrandomizer.com). The researcher shared the concealed group assignments with the study assessors via secured email. Randomisation to the nap or wake group was performed 1:1 for PD and HOA separately with block size 4 and was stratified for age (under 65 years vs. 65 years or older) and self-reported biological gender (male vs. female). The assessments were conducted at the Centre for Sleep Monitoring of the University Hospitals Leuven (UZ Leuven) in Belgium. The protocol was in accordance with the declaration of Helsinki and the World Medical Association, 2013 [38] and was approved by the Medical Ethical Committee Research UZ / KU Leuven (S61792). All participants provided written informed consent prior to study related procedures.

### Participants

Potential participants were recruited between November 2019 and December 2023 through a contact database of the Parkinson’s Disease Rehabilitation Research Group at the Department of Rehabilitation Sciences, KU Leuven, Belgium, or via a study flyer. Inclusion criteria comprised: (1) right-handedness, assessed with the Edinburgh Handedness Inventory [39]; (2) ≥40 years of age [40]; (3) Mini Mental State Examination (MMSE) score ≥24 [41]; (4) no history of comorbidities that could interfere with the study protocol. In addition to these criteria, people with PD were considered eligible if: (5) they had a diagnosis of PD made by a neurologist, according to the latest Movement Disorder Society diagnostic criteria [2] and (6) were in Hoehn & Yahr stage (H&Y) I-III ON medication [42]. Exclusion criteria for all participants were: (1) known diagnosis of insomnia; (2) severe apnoea (Apnoea Hypopnea Index (AHI) ≥30) as determined by an overnight screening polysomnography (PSG) [43]. PD-specific additional exclusion criteria were: (3) deep brain stimulation; (4) left-affected side for H&Y-I patients; (5) self-reported freezing of gait (FOG) with more than one freezing episode per month, according to the New Freezing of Gait Questionnaire [44].

The study was completed after successfully enrolling the anticipated sample size of 32 people with PD and 32 HOA, based on the current literature [20]. Participants were tested according to the randomised allocation (16 in the nap group and 16 in the wake group for both PD and HOA, see Figure 1). We maintained the hypotheses and protocol design according to the pre-registration, but we opted not to follow the outdated pre-registered analysis plan, to allow for the most direct comparison of the intervention effects. For this purpose, we used Bayesian statistics, complemented with inferential statistical tests, to aid the interpretation of the comparisons performed.

**Figure 1.**
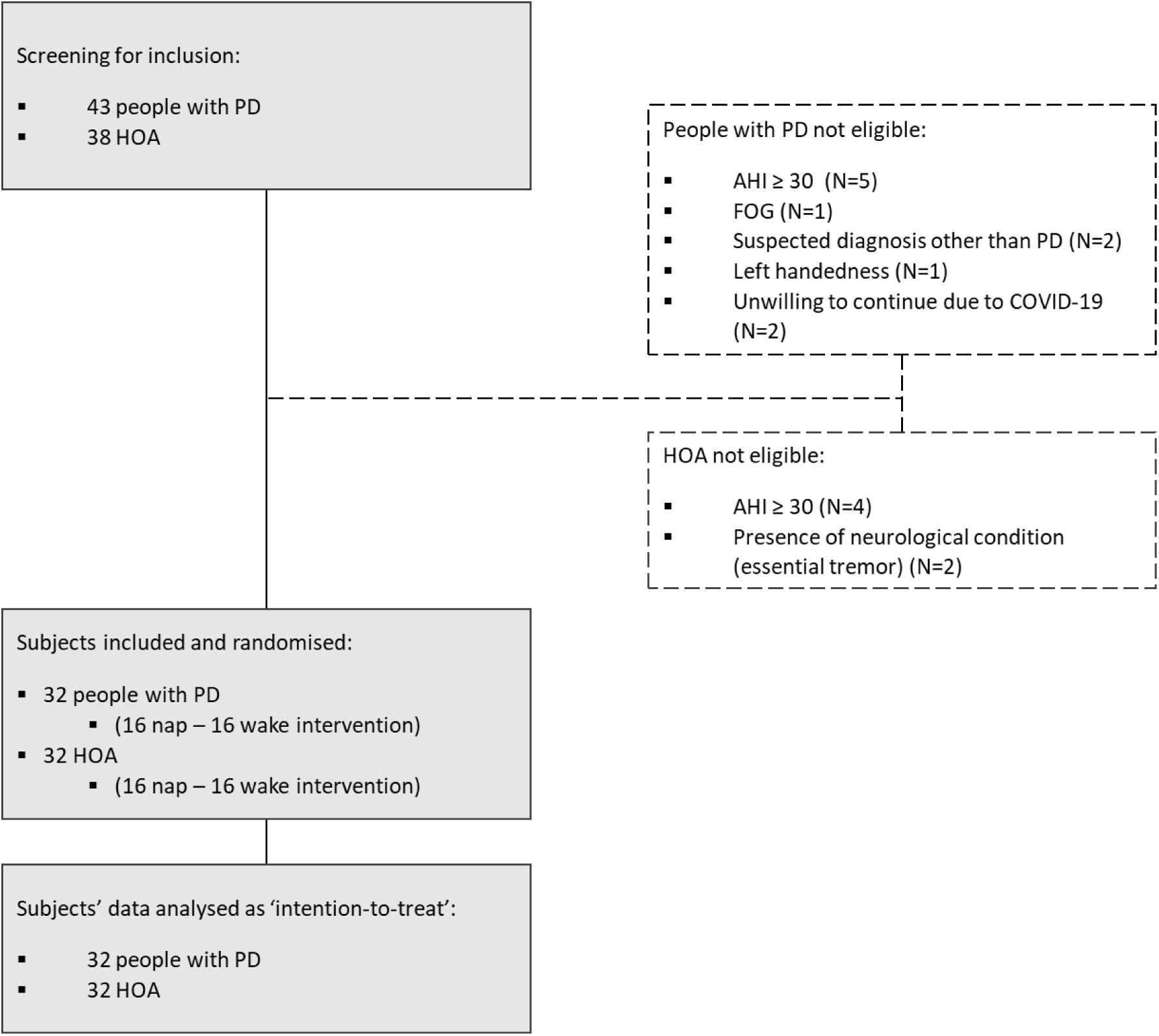
Study flow chart. A total of 43 people with Parkinson’s disease (PD) and 38 healthy older adults (HOA) were screened to reach the planned sample size of 32 enrolled participants per population. Five PD participants were not considered eligible due to an Apnoea Hypopnea Index (AHI) over 30 episodes per hour, 1 due toself-reported weekly Freezing of Gait (FOG), 1 for being left-handed according to the Edinburgh Handedness Inventory, 2 due to a presumed diagnosis other than PD (i.e., progressive supranuclear palsy, multiple systems atrophy), and 2 decided not to continue with the experiment after completion of screening due to health-related reasons unrelated to the study. Four HOA participants were not deemed eligible due to an AHI above 30, and 2 due to a comorbidity that could hamper the experimental procedures (essential tremor). There was no dropout after randomisation; all participants were assigned the randomised intervention, and there was no missing data on the primary and secondary outcomes, so all data was included in accordance to an intention to treat analysis.

### General Experimental Design

Prior to inclusion, a separate screening visit was scheduled. During this assessment, demographic information was obtained cognitive function was assessed with the Montreal Cognitive Assessment (MoCA) [45], and an overnight diagnostic PSG was performed to verify the AHI in all participants. The overnight PSG also served to familiarise participants to the experimental environment and EEG recording setup. People with PD were additionally tested on the Movement Disorders Society Unified Parkinson Disease Rating Scale (MDS-UPDRS) [46]. Information was also collected on disease duration and the Levodopa Equivalent Daily Dose (LEDD) [47] was calculated. Subjective sleep quality scales were also completed by all participants, comprising the Scales for Outcomes in Parkinson’s Disease (SCOPA)-sleep [48], Epworth Sleepiness Scale (ESS) [49], Pittsburgh Sleep Quality Index (PSQI) [50] and the REM-sleep Behaviour Disorder Screening Questionnaire (RBDSQ) [51]. For all these sleep scales, we report missing data from one HOA and 2 PD in the wake group, and an additional missing value for the SCOPA-sleep night and for the PSQI from one person with PD in the nap group. Following the screening night, participants’ habitual sleep and motion levels were monitored for 7 days and nights with an actigraphy watch (Philips Respironics Actiwatch-II®), but analyses on these data are not reported in this manuscript. Eligible participants were then invited to return to the university hospital for the experiment, which took place at least one week after the screening visit. They were instructed to keep a regular sleep/wake schedule three days before and during the experiment. They were additionally instructed to go to bed before midnight and refrain from caffeine and alcohol on the test days. People with PD continued taking their regular PD medications during the experiment, which were documented. Although intake times were not standardised, medication intake was systematically logged, but exact intake time was not noted for one patient post-intervention and for another post-night. Notably, two people with PD were drug-naïve.

The experiment consisted of three test sessions across two days (see Figure 2). At the beginning of each session, participants performed a Psychomotor Vigilance Test (PVT) of 100 stimuli to assess their level of vigilance [52] (see Supplementary Material 1 for details). Prior to initial practice on the first session (pre-intervention, ∼11 a.m. of Day 1), participants were tested on a motor execution task (MET, see section Motor Tasks below). This was followed by the testing of a unimanual self-initiated motor sequence learning (MSL) finger-tapping task first in a dual-task configuration, and then training and testing runs of the same sequence in a single-task configuration. After the pre-intervention session, participants were offered a standardised lunch, followed by a 2-hour opportunity to sleep or to remain awake quietly in supine or sitting position for an equivalent time period, as determined by the randomisation. Participants in the wake group were allowed to read and/or watch television, but they were not allowed to practise the MSL task or to do any form of physical activity. During the intervention period, all participants were monitored with low-density PSG, including those in the wake group, to supervise their wakeful state. Upon detection of drowsiness or N1 in the wake group, the assessor would enter the room to arouse the participant. The second session (post-intervention) started 30-40 minutes after the 2-hour nap/wake opportunity period to allow for possible sleep inertia effects to dissipate [53]. During the post-intervention session, the training and testing of the same sequence in a single-task configuration were again performed, and served as immediate post-intervention assessment. The single-task training and testing was followed by the dual-task testing. Lastly, a third session (post-night, on Day 2, 24 hours after the post-intervention session) comprised the training and testing of the sequence, first in a single-task configuration, then in dual-task. Notably, during the post-intervention and the post-night sessions, the single-task MSL (training and test) were performed first, followed by the dual-task MSL and MET to prevent interference of the dual-task practice on the consolidation assessment. Between Day 1 and Day 2, participants slept at home.

**Figure 2.**
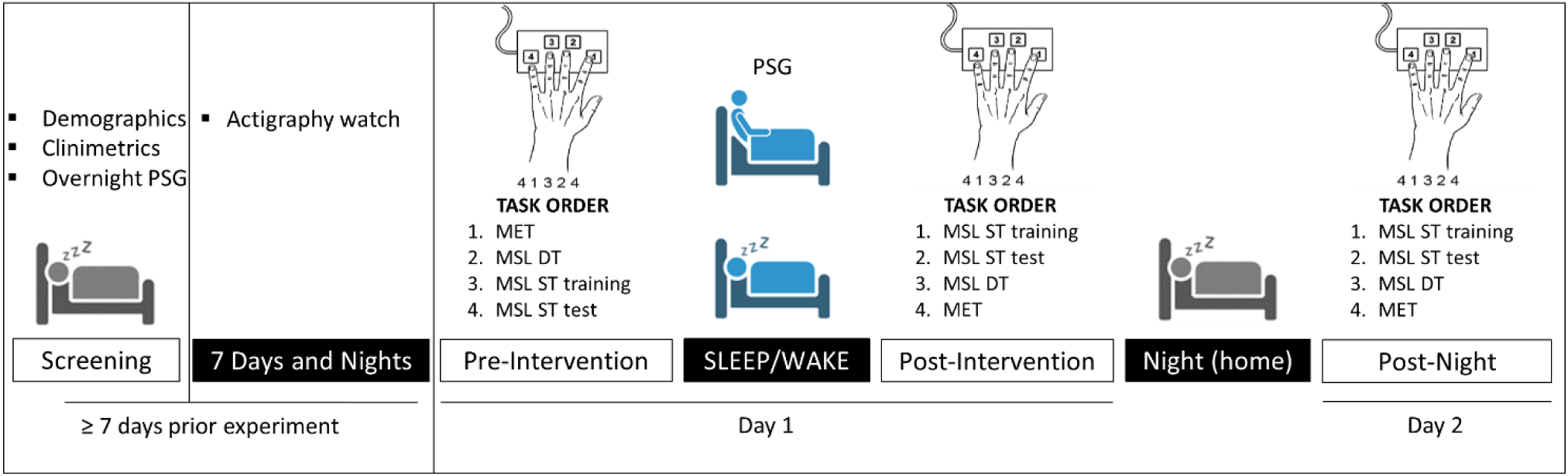
Experimental design with pre- and post-intervention assessments, as well as a 24-hour retention assessment (post-night). Note: MET = Motor Execution Test with four blocks of 48 key presses each sequence; DT = Dual-Task, with four blocks of 50 key presses sequence; ST, Single-Task training sequence, 14 blocks of 50 key presses; ST test, four blocks of 50 key presses sequence; PSG = polysomnography

### Motor tasks

All the tasks were scripted in Matlab (Math Works Inc, Natick, MA, USA; version R2018a), using the Psychophysics Toolbox extensions [54–56]. Participants were sitting comfortably in front of a laptop screen and used a left-handed Celeritas 5-Button Response Unit (Psychology Software Tools inc. Pittsburgh, USA) to practice the MET and the MSL with their left hand (non-dominant). The PVT was also performed in front of the laptop screen, but the spacebar of the keyboard was used as response button with the right (dominant) hand.

Both the MET and the MSL consisted of self-initiated unimanual finger-tapping tasks [14,57], during which the respective sequences were presented on the laptop screen for the whole duration of the practice blocks. During the MET, training and testing of the MSL, participants were instructed to perform the displayed sequence as quickly and as accurately as possible with their left, non-dominant, hand positioned on their lap under the table, to avoid visual feedback. No feedback on performance was provided during or after the training and testing. During these tasks, the timing of each key press and the corresponding response were recorded for data processing.

The MET consisted of four blocks of 48 key presses (ideally 12 correctly typed sequences). During each block, participants had to continuously type the sequence of numbers 4-3-2-1, with 4 and 1 corresponding to the little and index fingers, respectively. Each block was separated by a 20-second break, during which the sequence of numbers was concealed. This overtrained sequence served as a control condition to determine if any improvement observed on the MSL was due to sequence learning or to general improvement in motor execution of the task.

The MSL training consisted of 14 blocks of 50 key presses (ideally 10 correct sequences, i.e., ‘training’) with a 20-second break between each block [20]. During the 20-second rest period the sequence was concealed. After training, all participants had a 2-minute break to allow fatigue dissipation [58] and this was followed by another four practice blocks (i.e., ‘test’). The sequence of numbers for the MSL task was 4-1-3-2-4[14,57]. Before training, participants were first allowed to type this sequence three times slowly and as correctly as possible while looking at their hand to ensure they understood the task instruction.

The dual-task version of the MSL task consisted of four blocks of 50 key presses of the same MSL sequence, but in addition participants were instructed to count the number of substitutions of a cross sign (“+”) visible in the centre of the screen, below the sequence, by a circle sign (“o”) occurring at 5 to 8 pseudo-random moments during each block. Participants were instructed to keep count in their mind while typing the sequence as fast and accurately as possible without using a strategy (e.g., count aloud, keep the count with the fingers of the other hand). At the end of each block, they verbally reported the count to the assessor to verify that they were actively engaged in the secondary task. Their responses were documented but not further analysed as they do not allow for an assessment of a dual-task cost of the secondary task.

### Preprocessing

Sequence duration was calculated as the average time between the first and last key press of each fully correctly typed sequence (MSL = 4-1-2-3-4; MET = 4-3-2-1) in each block, and accuracy as the number of complete sequences typed correctly from the first to the last key, within a block. The primary dependent variable was the performance index (PI), a joint measure of sequence duration and accuracy on the MSL and MET tasks, derived for each block separately [20,25] (see Equation 1).

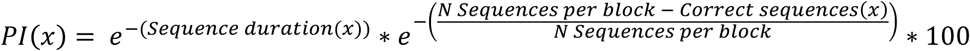

*Equation 1. Performance Index (PI) calculated per block (x). The sequence duration (x) consists of the mean sequence duration for each block, calculated on the correct sequences. The accuracy component is expressed as the ratio of correctly typed sequences by the total number of sequences per block (N = 10)*.

Sequence duration values above or below three standard deviations of the participant’s mean sequence duration of each block were considered outliers and subsequently excluded. This occurred only during the MET (MET PD = 0.93%; MET HOA = 0.87%), not during the MSL blocks.

### Polysomnography

During the screening night and the experimental intervention (nap/wake period), participants were monitored with PSG using the Medatec Brainnet system and Brainnet Software (Medatec, Brussels, Belgium, AASM2019 compliant).

For the screening night, EEG recordings comprised eight electrodes placed on the scalp (F3, F4, C3, C4, O1, O2, A1, A2), according to the international 10-20 system. A2 was used as recording reference. An electrode positioned in the centre of the forehead (Fpz), served as ground. Electrooculography (EOG) was recorded with two electrodes placed 1 cm from the external canthus of both eyes, below the left and above the right eye [59]. Electromyography (EMG) was recorded with two electrodes, on the chin and on the sublingual muscle. Additionally, two electrodes were placed on the lower limbs to screen for restless limbs syndrome. Another two electrodes were used to capture heart rate. Finally, two breathing bands, one around the chest and one around the abdomen, a sensor for oro-nasal pressure, sound and oxygen saturation completed the PSG setup.

During the experimental intervention a modified setup was used, comprising eight EEG (Fz, C3, Cz, C4, Pz, Oz, A1, A2), two EOG and two EMG (chin and sublingual muscle) electrodes, as well as the two breathing bands. Again, Fpz constituted the ground and A2 was used as recording reference. Impedances for the electrodes were kept below 10 kΩ [60]. PSG data were collected with a sampling frequency of 500 Hz.

### Preprocessing

For offline scoring of the naps, the electroencephalography (EEG) recordings were band-pass filtered (0.16-70 Hz) with Brainnet software (Medatec, Brussels, Belgium). These were then manually scored by an experienced sleep researcher, unblinded for the pathology, according to the AASM guidelines [61] on 30-second epochs using the fMRI Artefact rejection and Sleep scoring Toolbox (FASST) [62], implemented in MATLAB (Math Works Inc, Natick, MA, USA). We extracted percentage and duration of each sleep stage, total sleep time and sleep opportunity, and we calculated sleep efficiency as the percentage of time spent in sleep (e.g., N1, N2, N3 and REM) relative to the total time in bed.

As part of an explorative analysis, the sleep electrophysiological events occurring during the intervention were determined. To do so, epochs of N2-N3 sleep were extracted and further processed with FieldTrip Toolbox [63] in MATLAB. Raw data were resampled to 100 Hz and band-passed (0.5 – 30Hz) using Finite Impulse Response (FIR) filters. Data were subsequently manually screened for cleaning. Segments presenting abnormal muscular activity or eye movements were excluded. Next, Independent Component Analysis was performed, excluding components that presented cardiac artifacts. Electrophysiological markers of plasticity during sleep were detected using YASA toolbox [64] implemented in the Python environment. The YASA spindle detection algorithm was inspired by Lacourse and colleagues [65]. Specifically, the relative power in the spindle frequency range (10–16Hz) was calculated as a ratio to the total power in the broad-band range (1–30Hz) using Short-Time Fourier Transforms, applied with 2-second windows and a 200ms overlap. Next, the algorithm employed a 300ms window with a 100ms step size to calculate the moving root mean squared (RMS) of the EEG data filtered in the sigma band. A moving correlation was then calculated between the broadband signal (1–30Hz) and the EEG signal filtered in the spindle frequency band. Sleep spindles were detected upon the simultaneous reaching of three thresholds: (1) a relative power in the sigma band (with respect to total power) above 0.2; (2) a moving RMS crossing the RMS mean + 1.5 RMS standard deviation threshold and (3) moving correlation above 0.65. Additionally, the detected spindles with duration below 0.3s or above 3s were discarded. Spindles occurring on different channels within 500ms of each other were merged, as they were considered to represent the same event.

Slow wave detection was performed based on previous reports [66,67]. The signals were first bandpass filtered between 0.3–1.5Hz using a FIR filter. Negative peaks with an amplitude between –40 and –200μV and positive peaks with an amplitude comprised between 10–150μV were detected in the filtered signal. Next, the slow wave was composed of the negative peak and the nearest following positive peaks. A true slow wave was identified when the following criteria were met: (1) a duration of the negative deflection between 0.3–1.5 s, (2) a duration of the positive deflection between 0.1–1.0s, (3) a negative peak amplitude between 40–200 μV, (4) a positive peak amplitude between 10–150μV and (5) a peak-to-peak amplitude between 75– 350 μV.

Events were identified in all electrodes, but only spindles detected in C3 and slow waves detected in Fz were further analysed. The amplitude and frequency of each detected spindle and the amplitude and slope of each detected slow wave were extracted [67,68]. We then computed the phase-amplitude coupling between the phase of slow waves (0.3–1.5 Hz) and the spindles-related sigma band amplitude (10–16 Hz). To do so, the normalised direct phase-amplitude coupling (ndPAC) was computed at the highest sigma peak amplitude within a 2 second epoch centred around the current slow-wave trough. The ndPAC allows to quantify the strength of the phase amplitude coupling [69–72].

We also extracted spindle and slow wave density, defined as the number of these events occurring per minute of NREM2 and NREM3 sleep. Features of spindles, slow waves and ndPAC above and below 3 standard deviations of each participant’s mean were considered outliers (spindle amplitude: PD = 1.18%, HOA = 1.10%; spindle frequency: PD = 0.09%, HOA = 0.19%; slow wave amplitude: PD = 1.47%, HOA = 1.59%; slow wave slope: PD = 1.00%, HOA = 1.10%; ndPAC: PD = 0.03%, HOA = 0.02%), and were therefore excluded from further analyses. Overall, we succeeded in the detection of spindles both in PD and in HOA (PD: mean = 345.1, range [45–877]; HOA: mean = 318.2, range [60–553]), as well as of slow waves (PD: mean = 191.3, range [6–1020]; HOA: mean = 131.9, range [1–411]). In one HOA, the algorithm did not detect slow waves in the channel Fz, but four slow waves were detected in channels C3, Cz, Pz and Oz.

### Statistical analyses

Statistical analyses were performed for PI, sequence duration and accuracy. As the PI was our primary metric of interest, we report these results in the main text. Findings on sequence duration and accuracy are detailed in Supplementary Materials 3 and 4, respectively.

Analyses were conducted with R open-source software [73]. For this phase-two explorative study we adopted the use of Bayesian statistical tests, indicating the likelihood that the observed data supported the acceptance of the alternative hypothesis (e.g., the two conditions have different means/variances) or the null hypothesis (e.g., the two conditions have similar means/variances). Frequentist statistics are also provided in this manuscript to allow for comparison with previous studies. We calculated Bayes factors (BF) to provide a clearer interpretation to the statistical comparisons, using the BayesFactor package [74]. Specifically, we present BF_10_ as a relative measure of evidence in favour of a difference, in comparison to the null hypothesis. To assess the strength of evidence against the null hypothesis, we classified BFs according to the guidelines provided by Lee and Wagenmakers [75], where values above 1 indicate evidence in favour of the alternative hypothesis and values below 1 suggest evidence in favour of the null hypothesis. Evidence was reported in different degrees (1 < BF ≤ 3 as weak evidence, 3 < BF ≤ 10 as moderate evidence, 10 < BF ≤ 100 as strong evidence, BF > 100 as decisive evidence). BFs below 1 indicated evidence in favour of the null hypothesis in different degrees (0.33 ≤ BF < 1 as weak evidence, 0.01 ≤ BF < 0.33 as moderate evidence, 0.001 ≤ BF < 0.01 as strong evidence and BF < 0.001 as decisive evidence) [75]. Evidence defined as weak (0.33 ≤ BF < 3) should be regarded as inconclusive and may be related to a lack of statistical power. For the analyses using Bayesian statistics, (i) we assigned a noninformative Jeffreys prior for the variance and a Cauchy prior for the standardised effect size with value √2/2, (ii) implemented 10.000 Markov Chain Monte Carlo iterations, and (iii) we excluded the non-available values. Post-hoc comparisons were performed when BFs were above 3 (i.e., moderate evidence in favour of the rejection of the null hypothesis).

Bayesian analyses of variance (ANOVA) or general Bayesian tests were used to obtain BFs using the functions “anovaBF” and “generalTestBF” from the R package BayesFactor [74]. For the Bayesian ANOVA’s, the null model included only the random effect, while for the general Bayesian tests the null model comprised the intercept. We set the parameter “whichModels” to “all” in order to extract the appropriate BFs. We kept the default values for the prior probability of the fixed and random effects. For reference, these analyses were also performed with frequentist statistics, using repeated measures ANOVA’s with random effect of subject or two-way ANOVA’s, implemented in the ez package in R [76] (Fisher’s F statistics and p-values are reported). Greenhouse-Geisser (GG) correction was applied in case of violation of the sphericity assumption.

Post-hoc tests were performed using Bayesian t-tests. For these, the null model held the assumption that the difference between the means is zero, and we calculated BFs using the function “ttestBF” [74]. We complemented these Bayesian statistical analyses with frequentist Student’s t-tests or Mann-Whitney U tests and we report the confidence interval for the mean difference. For Bayesian correlation analyses, we reported the BFs output by the function “correlationBF” from the package BayesFactor [74]. Frequentist correlation analyses were performed with Pearson’s r tests or Spearman r tests (r and p-values are reported). For comparing the correlations obtained, r-scores from PD and HOA data were firstly transformed to Z values using the Fisher-z transformation [77], and were then compared using the function “cocor.indep.groups” implemented in the cocor package [78], with alpha = 0.05 and confidence intervals reported as per Zou [79]. In case the data of one group was not normally distributed, Spearman correlations were calculated on both groups and then compared.

Additionally, effect sizes were reported as generalised eta squared (ges) [80], Cohen’s d (for frequentist parametric tests) or rank-biserial correlation (r_rb_) for frequentist non-parametric tests. The r_rb_ was computed as the Spearman correlation coefficient between a dichotomous variable (e.g., group, intervention) and the outcome of interest. Negative values for the effect sizes favour the PD group in comparison to HOA. When comparing interventions, positive values suggest an effect favouring the nap over the wake intervention. Values of r_rb_ should be interpreted according to the values reported by Cohen, 1992, and are complemented with the p-value of the correlation, to indicate their significance [81].

### Behaviour

#### Pre-requisites

MSL performance was tested against that of the MET, to verify whether the differences observed over the three sessions were related to sequence-specific learning or to general motor execution improvement. As the MET sequence consisted of four key presses, in contrast with the five of the MSL sequence, only relative performance changes were compared. Specifically, the relative difference between the MET four blocks pre-intervention and post-night, was compared to the relative difference between the MSL first four blocks of single-task training pre-nap and the last four blocks single-task test post-night. For this analysis, a Bayesian ANOVA was used, including group, intervention and task (MSL/MET) as fixed effects, and subject as random effect, against the null model including only the random effect (subject). Overall, these results suggest that the performance change was not related to general motor execution improvement only, but to actual learning, irrespective of group and intervention (results in Supplementary Material 2).

Mean values of the fourteen blocks of training pre-intervention were used to verify that the MSL sequence was successfully encoded. For this analysis, we used a Bayesian ANOVA including group, intervention and block (14), and subject as random effect, and reported the BFs for each of these variables when compared to the null model. Notably, three people with PD (two allocated to the nap, one to the wake intervention) performed no correct MSL sequence for a total of seven missing datapoints, which were excluded from the analysis.

To test whether performance reached a plateau before the intervention we explored the differences in PI among the four blocks of test pre-intervention. Here, a first block effect was found (results in Supplementary Material 2), therefore we excluded this first block of test pre-intervention and the first block of training post-intervention in the calculation of offline performance changes post-intervention, both in PD and in HOA, as previously done in another study on sleep and memory consolidation in young healthy participants [82]. Similarly, the first block was removed from the test blocks post-intervention and from the first four blocks of training post-night for the calculation of post-night offline relative changes (Supplementary Material 2).

To verify that medication did not influence learning and memory consolidation in our study, we performed a control explorative analysis where we correlated the post-intervention and post-night offline changes, as well as dual-task costs (see below) post-intervention and post-night, with the levodopa equivalent daily dose (LEDD) for the nap and wake group separately. We found weak evidence in favour of no correlation between the two variables (see Supplementary Material 5). LEDD was thus not included as a confounder in the analyses of offline changes.

#### Main analyses

We tested for the effects and the interactions of the intervention (nap/wake) and the group (PD/HOA) using Bayesian ANOVA’s. Our primary interest was the relative offline change in PI between the mean of the last three blocks of test pre-intervention and that of blocks 2-4 of single-task training post-intervention. The secondary interest were the overnight changes in performance, measured as the performance change between the last three blocks of test post-intervention and the first three blocks, following the very first, of single-task training post-night. Again, we compared the two interventions in the PD and HOA group using Bayesian ANOVA’s.

Automaticity was assessed as the dual-task cost computed as the relative difference between dual-task performance and single-task performance at post-training test, both at post-intervention and at post-night separately. These values were compared using Bayesian ANOVA’s including intervention (nap/wake) and group (PD/HOA) and their interaction. For this analysis we did not exclude the first block of test single-task or of dual-task MSL, since the reaching of a performance plateau was not a requirement for the computation of dual-task cost [83,84].

In agreement with current evidence suggesting that sleep not only impacts consolidation, but may also have a beneficial effect on extended practice [20,23], we also tested whether napping had an effect on performance changes between the beginning and the end of the training (extended practice) post-intervention and post-night. For this analysis we employed Bayesian ANOVA’s (group x intervention). For each session (i.e., post-intervention and post-night), the relative difference of the mean of the first four blocks (start) and that of the last four blocks (end) of MSL training was calculated.

### Electroencephalography

The electrophysiological sleep parameters obtained during the experimental nap were compared between people with PD and HOA using general Bayesian general tests (“generalTestBF”), supplemented by Analyses of Covariance (ANCOVAs), implemented using the “lm” function from the stats package [73]. In these analyses, we included gender, age, and AHI, as they are known to influence sleep micro-architecture [85] (results in Supplementary Material 6). These covariates were added together with their interactions, to the group factor, and their effects compared to the null model (including only the intercept). The main effects of the group for these analyses are reported in the main text, while the effects with highest BF are described in Supplementary Material 6. Additionally, we tested the correlation between amount of N2-N3 sleep, characteristics of spindles (amplitude, frequency, density) and slow waves (amplitude, slope, density) as well as their coupling (ndPAC), and the relative offline change in MSL performance post-intervention using Bayesian correlations. These correlation coefficients were then compared, to highlight differences in the associations of sleep macro- and micro-architecture and performance changes.

## Results

The demographic characteristics of the included participants can be found in Table 1. According to the reported BFs, we found weak evidence supporting similarity between nap/wake intervention groups for age, gender, AHI, cognition (measured with MoCA), sustained attention (measured with PVT) and subjective sleep scales, within each population group (PD/HOA). Also, weak evidence was found for higher sustained attention pre-compared to post-intervention in the people with PD allocated to the nap group (BF_10_ = 1.29, V= 106, p = 0.05), but this was weakly to moderately in favour of no difference for the other comparisons between pre- and post-intervention (PD-wake: BF_10_ = 0.35, V = 84, p = 0.43; HOA-nap: BF_10_ = 0.26, V = 90, p = 0.27; HOA-wake: BF_10_ = 0.27, V = 57, p = 0.60). The analyses of PVT further showed higher sustained attention at post-intervention compared to post-night in people with PD and in HOA in the nap group (PD: BF_10_ = 1.07, V = 28, p = 0.04; HOA: BF_10_ = 2.35, V= 112, p = 0.02), but these were weakly favouring no difference in those allocated to wake (PD: BF_10_ = 0.47, V = 78, p = 0.63; HOA: BF_10_ = 0.82, t_(15)_ = 1.69, p = 0.11).

**Table 1.** General demographic characteristics.

|  | PD Nap<br>(=16) | PD Wake<br>(=16) | Test result<br>(PD<br>nap – wake) | HOA Nap<br>(=16) | HOA Wake<br>(=16) | Test result<br>(HOA<br>nap – wake) | Test result<br>(PD – HOA) |
| --- | --- | --- | --- | --- | --- | --- | --- |
| **Demographic statistics** |  |  |  |  |  |  |  |
| **Age**<br>**(years)**<br>**mean [range]** | 66.9<br>[53–78] | 65.7<br>[47–80] | $BF_{10} = 0.37$<br>$t_{(30)} = 0.47$<br>$p = 0.64$ | 65.56<br>[48–75] | 68<br>[51–78] | $BF_{10} = 0.49$<br>$t_{(30)} = -0.99$<br>$p = 0.33$ | $BF_{10} = 0.26$<br>$t_{(62)} = -0.26$<br>$p = 0.80$ |
| **Gender (M/F)** | 9/7 | 11/5 | | 11/5 | 11/5 | | $\chi^2_{(3)} = 0.44$<br>$p = 0.93$ |
| **AHI (n./hour)** | 9.1<br>[5.0–13.1] | 6.15<br>[2.9–9.4] | BF<sub>10</sub> = 0.58<br>W = 164.5<br>p = 0.17 | 11.2<br>[6.8–15.6] | 13.3<br>[8.6–18.0] | BF<sub>10</sub> = 0.40<br>W = 115<br>p = 0.64 | BF<sub>10</sub> = 2.84<br>W = 333<br>p = 0.01 |
| **MoCA (0–30)** | 26.26<br>[25.1–27.4] | 27.13<br>[25.7–28.6] | BF<sub>10</sub> = 0.49<br>W = 89<br>p = 0.14 | 26.56<br>[25.5–27.6] | 27.31<br>[26.2–28.5] | BF<sub>10</sub> = 0.51<br>t<sub>(30)</sub> = -1.03<br>p = 0.31 | BF<sub>10</sub> = 0.28<br>W = 506<br>p = 0.94 |
| **PVT pre-intervention** | 0.36<br>[0.31–0.40] | 0.36<br>[0.30–0.41] | BF<sub>10</sub> = 0.34<br>W = 130<br>p = 0.96 | 0.35<br>[0.33–0.37] | 0.37<br>[0.34–0.39] | BF<sub>10</sub> = 0.52<br>W = 108.5<br>p = 0.47 | BF<sub>10</sub> = 0.26<br>W = 583<br>p = 0.34 |
| **PVT post-intervention** | 0.34<br>[0.31–0.38] | 0.38<br>[0.33–0.43] | BF<sub>10</sub> = 0.55<br>W = 126<br>p = 0.96 | 0.35<br>[0.32–0.37] | 0.37<br>[0.35–0.39] | BF<sub>10</sub> = 0.63<br>W = 88.5<br>p = 0.14 | BF<sub>10</sub> = 0.26<br>W = 528<br>p = 0.84 |
| **PVT post-night** | 0.39<br>[0.35–0.44] | 0.36<br>[0.33–0.40] | BF<sub>10</sub> = 0.48<br>W = 160<br>p = 0.24 | 0.34<br>[0.32–0.36] | 0.36<br>[0.34–0.38] | BF<sub>10</sub> = 0.84<br>W = 90.5<br>p = 0.16 | BF<sub>10</sub> = 1.48<br>W = 604<br>p = 0.22 |
| **PD-specific measures** |  |  |  |  |  |  |  |
| **Disease duration (years)** | 5.4<br>[3.6–7.2] | 5.2<br>[3.0–7.5] | BF<sub>10</sub> = 0.34<br>W = 141<br>p = 0.64 | - | - | - | - |
| **MDS-UPDRS III (0–132)** | 29.2<br>[23.6–34.8] | 31.6<br>[25.8–37.4] | BF<sub>10</sub> = 0.39<br>W = 109<br>p = 0.48 | - | - | - | - |
| **LEDD (mg/24h)** | 450.7<br>[323.4–578.0] | 656.4<br>[404.2–908.7] | BF<sub>10</sub> = 0.83<br>W = 94.5<br>p = 0.21 | - | - | - | - |
| **Dominant symptom side<sup>§</sup> (R/L)** | 9/7 | 9/7 |  | - | - | - | - |
| **Subjective sleep measures** |  |  |  |  |  |  |  |
| **PSQI (0–21)** | 7.2<br>[4.6–9.8] | 7.4<br>[5.3–9.5] | BF<sub>10</sub> = 0.35<br>W = 99.5<br>p = 0.83 | 3.4<br>[2.1–4.7] | 4.9<br>[3.5–6.3] | BF<sub>10</sub> = 1.00<br>W = 72<br>p = 0.06 | BF<sub>10</sub> = 26.59<br>W = 658.5<br>p < 0.01* |
| **SCOPA-sleep night (0–21)** | 5.3<br>[3.7–7.4] | 6.3<br>[4.1–8.5] | BF<sub>10</sub> = 0.39<br>t<sub>(27)</sub> = -0.53<br>p = 0.60 | 4.4<br>[2.5–6.3] | 3.0<br>[1.2–4.8] | BF<sub>10</sub> = 0.54<br>W = 151<br>p = 0.22 | BF<sub>10</sub> = 2.50<br>W = 617<br>p = 0.01* |
| **SCOPA-sleep day (0–18)** | 5.8<br>[3.9–7.6] | 7.5<br>[5.1–9.9] | BF<sub>10</sub> = 0.60<br>t<sub>(28)</sub> = -1.21<br>p = 0.24 | 2.75<br>[1.77–3.73] | 2.7<br>[1.4–3.9] | BF<sub>10</sub> = 0.34<br>t<sub>(29)</sub> = 0.11<br>p = 0.91 | BF<sub>10</sub> = 1287.79<br>W = 752<br>p < 0.01* |
| **ESS (0–24)** | 10.0<br>[7.6–12.4] | 10.6<br>[8.1–13.1] | BF<sub>10</sub> = 0.36<br>t<sub>(28)</sub> = -0.34<br>p = 0.74 | 4.9<br>[3.0–6.8] | 6.8<br>[4.2–9.4] | BF<sub>10</sub> = 0.59<br>W = 92<br>p = 0.27 | BF<sub>10</sub> = 97.79<br>W = 708<br>p < 0.01* |
| **RBDSQ (0–13)** | 4.6<br>[3.0–6.1] | 5.7<br>[4.2–7.2] | BF<sub>10</sub> = 0.54<br>W = 83<br>p = 0.23 | 2.9<br>[1.3–4.5] | 2.5<br>[0.8–4.3] | BF<sub>10</sub> = 0.39<br>W = 142<br>p = 0.38 | BF<sub>10</sub> = 601.69<br>W = 731.5<br>p < 0.01* |

AHI, Apnea Hypopnea Index; MMSE, Mini-Mental State Examination; MoCA, Montreal Cognitive Assessment; PVT, Psychomotor Vigilance Test; LEDD, Levodopa Equivalent Daily Dose; MDS-UPDRS, Movement Disorders Society – Unified Parkinson's Disease Rating Scale; PSQI, Pittsburgh Sleepiness Questionnaire, International; ESS, Epworth Sleepiness Scale; SCOPA-Sleep, Scales for Outcomes in Parkinson's Disease – Sleep; RBDSQ, REM sleep Behaviour Disorder Screening Questionnaire; HADS, Hospital Anxiety and Depression Scale.
§ The dominant symptom side was determined as the side with the higher sum of scores from the left-side and right-side items on the Movement Disorders Society – Unified Parkinson's Disease Rating Scale part III [86].
Variables are presented as the mean ( ± 95% confidence interval), unless otherwise specified
\* significant p values according to frequentist statistics

When comparing the two population groups irrespective of the intervention, weak evidence for a higher AHI for HOA than for PD was found (BF_10_ = 2.84). Evidence was decisively in favour of a true difference for the RBDSQ, which was higher in PD (BF_10_ = 601.69). Strong to decisive evidence was found for worse daytime sleepiness in PD compared to HOA, as measured with the ESS (BF_10_ = 97.79) and the SCOPA-Sleep Day (BF_10_ > 1000). For night-time sleep quality, there was strong evidence in favour of a difference for the PSQI (BF_10_ = 26.59), but this was only weakly in favour of a difference between the two groups for the SCOPA-Sleep night (BF_10_ = 2.50). These findings suggest that the two populations were similar, except for subjective sleep scales, where people with PD reported worse sleep quality as compared to HOA (see Table 1).

### Behavioural data

Figure 3 shows the overall performance of the PD (Figure 3A) and HOA (Figure 3B) groups across the three sessions, measured with the PI.

**Figure 3.**
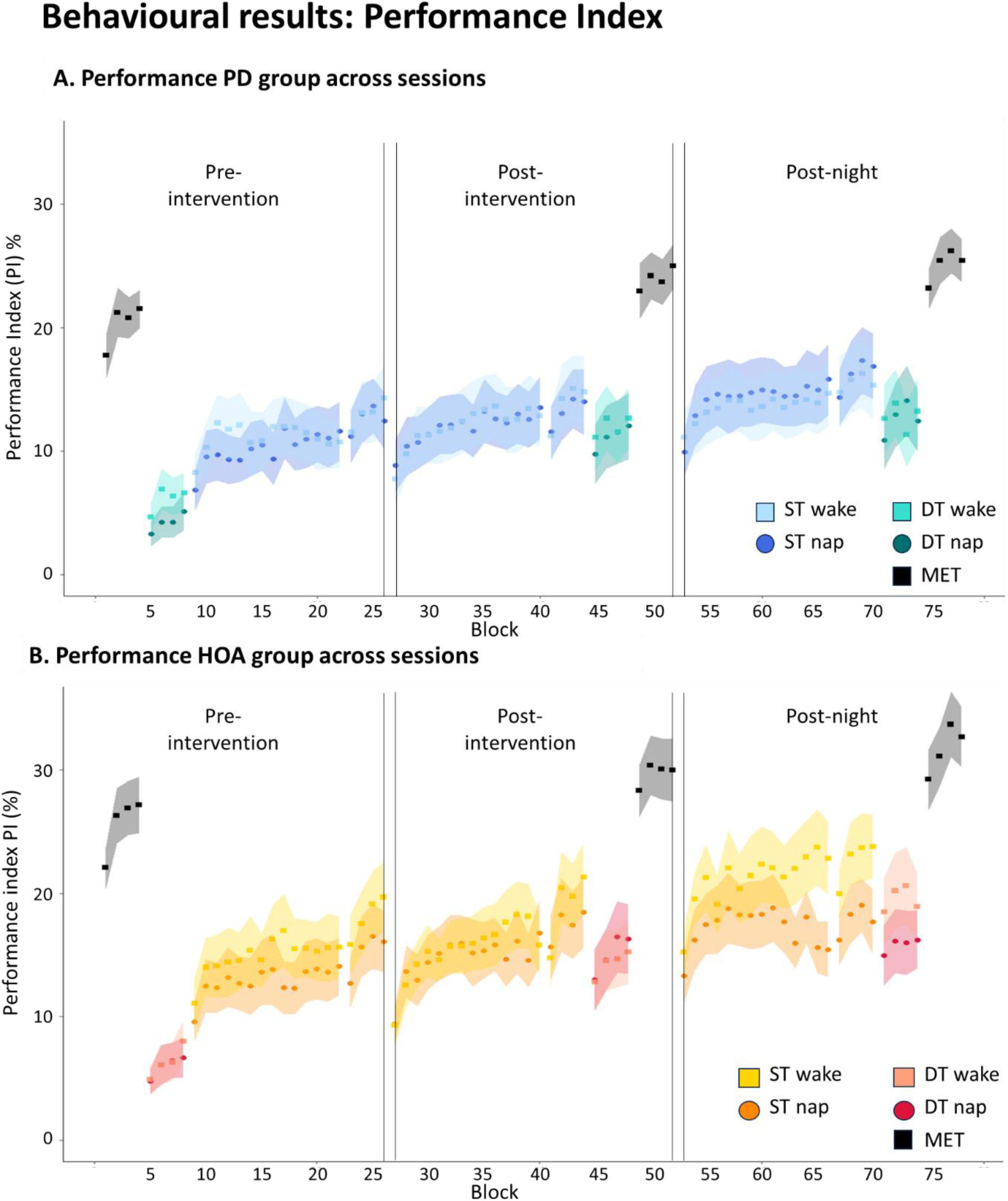
Representation of the performance index (PI) over the three sessions, expressed as mean and standard error (A) for the PD group (in cold colours) and (B) for the HOA (in warm colours). MET: motor execution test; ST: single task; DT: dual-task. Higher values indicate better performance.

#### Initial learning

First, we tested whether the MSL sequence was successfully encoded. Findings from the Bayesian ANOVA showed decisive evidence for an effect of block (BF_10_ > 1000, F_(13,741)_ = 7.66, p < 0.01, p(GG) < 0.01, ges = 0.01), with an overall trend for improvement during the training, but weak to strong evidence for no effect of all the other main factors and interactions (group: BF_10_ = 0.68, F_(1,57)_ = 1.20, p = 0.28, ges = 0.02; intervention: BF_10_ = 0.54, F_(1,57)_ = 0.31, p = 0.58, ges < 0.01; group by intervention: BF_10_ = 0.64, F_(1,57)_ = 0.15, p = 0.70, ges < 0.01; group by block: BF_10_ = 0.001, F_(13,741)_ = 0.38, p = 0.98, p(GG) = 0.90, ges < 0.01; intervention by block: BF_10_ = 0.004, F_(13,741)_ = 0.83, p = 0.63, p(GG) = 0.56, ges < 0.01; group by intervention by block: BF_10_ = 0.04, F_(13,741)_ = 1.13, p = 0.33, p(GG) = 0.34, ges < 0.01). These results suggest that learning of the MSL occurred over time, but it was not different between populations and intervention groups.

#### Offline MSL changes post-intervention

Our primary interest was the consolidation post-intervention of the MSL single-task. We found overall weak evidence for no difference between groups and interventions (group: BF_10_ = 0.49, F_(1,60)_ = 1.54, p = 0.22, ges = 0.03; intervention: BF_10_ = 0.51, F_(1,60)_ = 1.63, p = 0.21, ges = 0.03), nor for an interaction effect (BF_10_ = 0.41, F_(1,60)_ = 1.12, p = 0.29, ges = 0.02) (Figure 4A). These findings suggest no differential effect of a period of post-learning sleep compared to an equivalent time awake on motor memory consolidation, neither in PD nor in HOA, when measured with PI.

**Figure 4.**
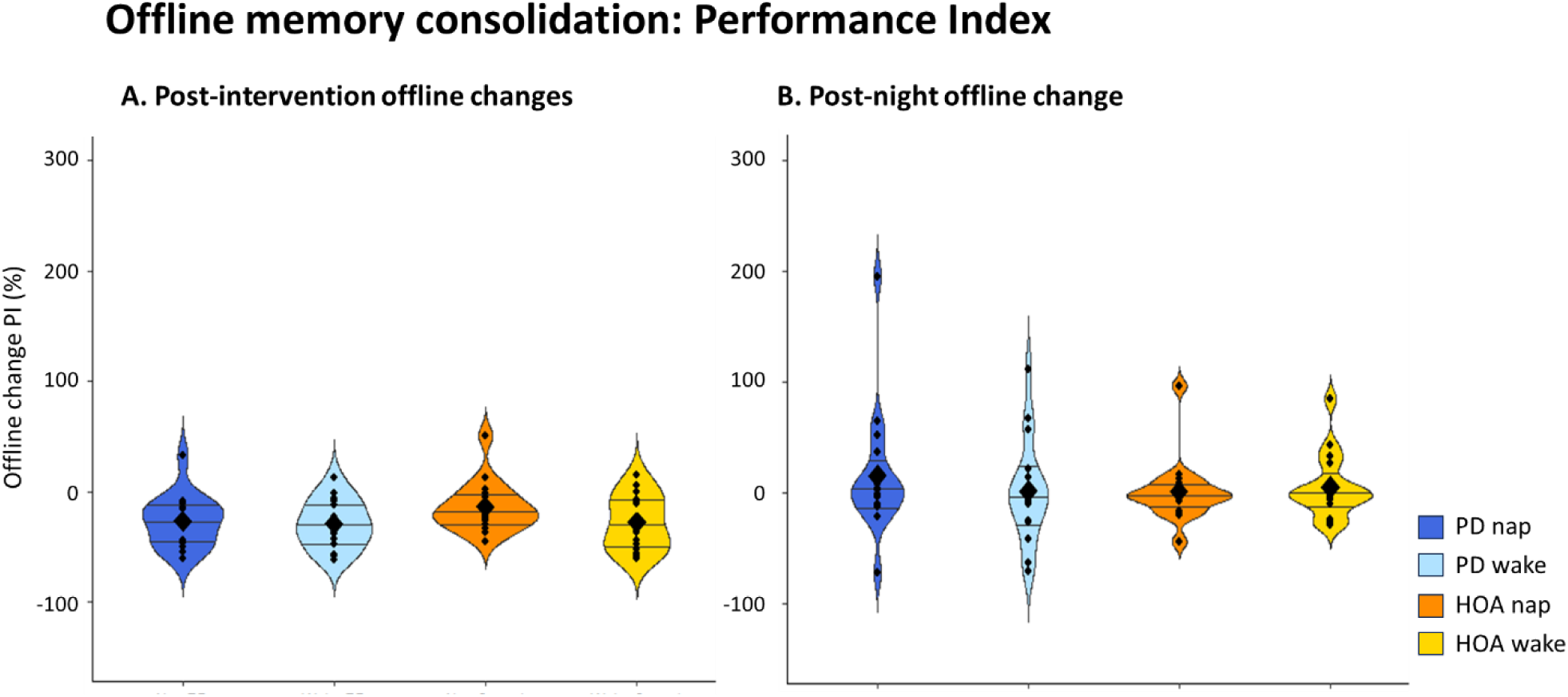
(A) Post-intervention offline changes of single-task MSL performance in the PD and HOA groups. (B) Post-night offline changes of single-task MSL performance in the PD and HOA groups. Violin plot: mean (diamond), median (central horizontal bar), and 25^th^ (lower bar) and 75^th^ (higher bar) percentiles.

#### Offline MSL changes post-night

Our secondary interest was the post-night performance change of the MSL sequence. The analyses revealed weak to moderate evidence for no effect of group, intervention (group: BF_10_ = 0.38, F_(1,60)_ = 0.89, p = 0.35, ges = 0.01; intervention: BF_10_ = 0.32, F_(1,60)_ = 0.49, p = 0.49, ges < 0.01) and their interaction (BF_10_ = 0.32, F_(1,60)_ = 0.54, p = 0.46, ges < 0.01) (Figure 4B). These results indicate that the sleep intervention did not generate protracted benefits after 24 hours, and offline changes at post-night were similar between PD and HOA.

#### Dual-task costs

Automaticity of the MSL task was tested with the dual-task cost, whereby higher values indicate larger interference of the secondary task. At post-intervention, the analyses showed weak to moderate evidence for no effect of group (BF_10_ = 0.28, F_(1,60)_ = 0.23, p = 0.63, ges < 0.01), intervention (BF_10_ = 0.45, F_(1,60)_ = 1.33, p = 0.25, ges = 0.02) or their interaction (BF_10_ = 0.45, F_(1,60)_ = 1.36, p = 0.25, ges = 0.02) (Figure 5A). Similar results were found at post-night (group: BF_10_ = 0.69, F_(1,60)_ = 2.32, p = 0.13, ges = 0.04; intervention: BF_10_ = 0.26, F_(1,60)_ = 0.06, p = 0.81, ges < 0.01; group by intervention: BF_10_ = 0.42, F_(1,60)_ = 1.84, p = 0.28, ges = 0.02) (Figure 5B). These findings suggest that the interventions had similar effects on automaticity in PD and in HOA, both in the short- and in the longer-term.

**Figure 5.**
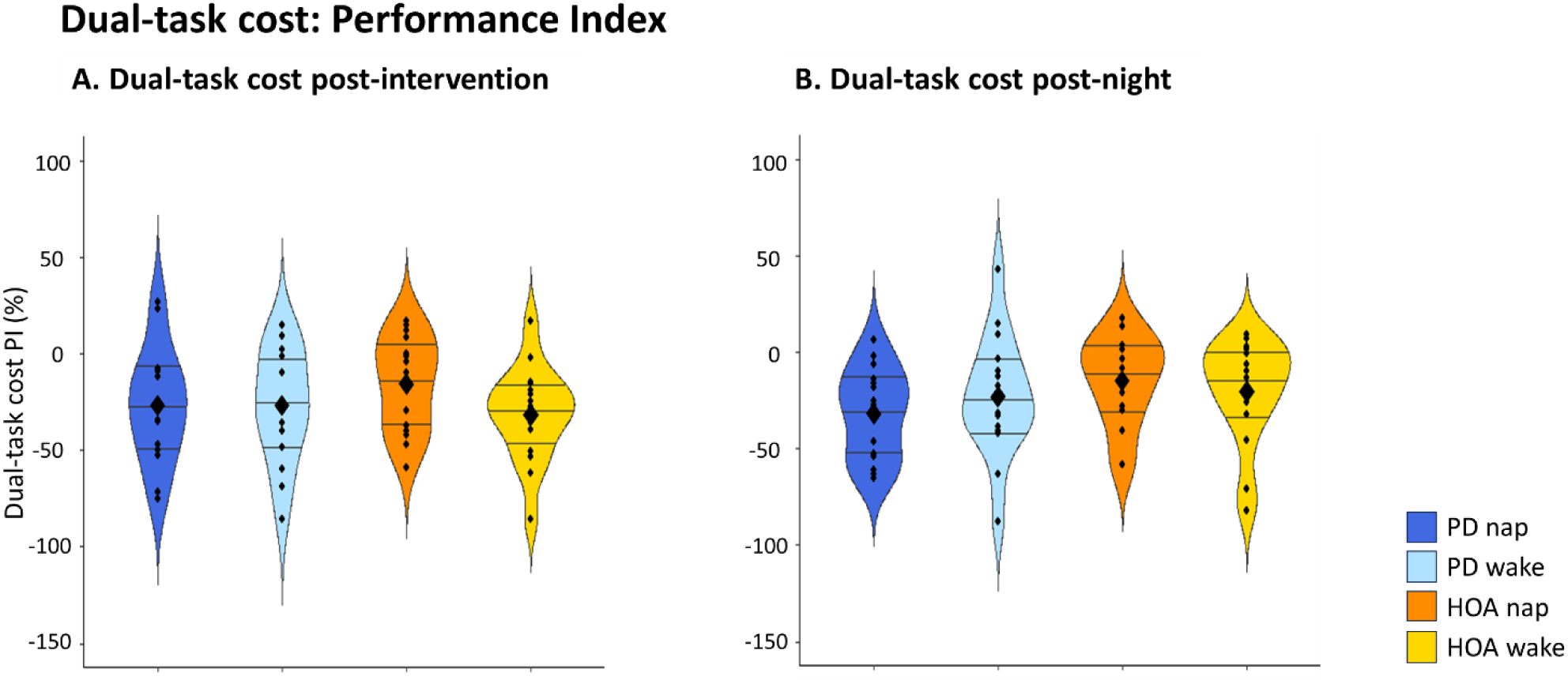
Dual-task costs. (A) Post-intervention dual-task cost of performance of the MSL sequence in the PD and HOA groups. (B) Post-night dual-task cost of performance of the MSL sequence in the PD and HOA groups. Violin plot: mean (diamond), median (central horizontal bar), and 25^th^ (lower bar) and 75^th^ (higher bar) percentiles.

#### Extended practice post-intervention and post-night

At post-intervention, weak to moderate evidence was found for no effect of group (BF_10_ = 0.69, F_(1,60)_ = 2.28, p = 0.14, ges = 0.04), intervention (BF_10_ = 0.27, F_(1,60)_ = 0.16, p = 0.69, ges < 0.01), and group by intervention interaction (BF_10_ = 0.27, F_(1,60)_ = 0.16, p = 0.69, ges < 0.01). At post-night, the two-way ANOVA provided weak evidence for a group effect favouring the HOA (BF_10_ = 1.15, F_(1,60)_ = 3.69, p = 0.06, ges = 0.06), and weak evidence for no effect of intervention (BF_10_ = 0.41, F_(1,60)_ = 1.21, p = 0.28, ges = 0.02), nor group by intervention interaction (BF_10_ = 0.61, F_(1,60)_ = 2.18, p = 0.15, ges = 0.04).

Hence, the rate of performance change during training was not different between PD and HOA post-intervention. However, the results weakly suggest a possible group difference post-night. Notably, the intervention appeared to have no effect on extended practice measured with PI at post-intervention, but better performance changes were noted in the HOA at post-night.

### EEG analyses

Bayesian t-tests revealed weak evidence for no difference between people with PD and HOA on overall sleep efficiency, percentages of N1, N2, N3, and REM (see Table 2). All participants presented at least 14 minutes of N2 sleep. Moreover, twelve HOA presented N3 sleep, and eight also showed REM sleep. In the PD group, 10 of the 16 participants allocated to the nap intervention showed N3 sleep and three reached REM sleep.

**Table 2.** Sleep macro-architecture metrics from the nap in PD and HOA.

|  | PD (= 16) | HOA (= 16) | Test result<br>(PD – HOA) |
| --- | --- | --- | --- |
| Overall sleep efficiency (%) | 54.8 [42.0 – 67.5] | 48.8 [37.2 – 60.4] | $BF_{10} = 0.40$<br>$t_{(30)} = 0.68$, $p = 0.50$ |
| N1 (%) | 11.8 [8.7 – 14.8] | 14.2 [8.8 – 19.6] | $BF_{10} = 0.42$<br>$W = 125$, $p = 0.93$ |
| N2 (%) | 41.6 [33.0 – 50.2] | 34.5 [27.6 – 41.5] | BF<sub>10</sub> = 0.61<br>t<sub>(30)</sub> = 1.25, p = 0.22 |
| N3 (%) | 11.3 [3.4 – 19.2] | 11.9 [5.5 – 18.3] | BF<sub>10</sub> = 0.34<br>W = 117, p = 0.69 |
| REM (%) | 1.8 [-0.5 – 4.1] | 2.4 [0.9 – 3.9] | BF<sub>10</sub> = 0.36<br>W = 92, p = 0.11 |

*Sleep efficiency: percentage of total sleep time/total time in bed. The values are reported as mean [ ± 95% confidence interval]*

Comparisons between PD and HOA on electrophysiological markers of plasticity during sleep revealed overall weak evidence for no group difference (spindle amplitude: BF_10_ = 0.89, F_(1)_ = 0.77, p = 0.39, ges = 0.02; spindle frequency: BF_10_ = 0.34, F_(1)_ = 0.03, p = 0.87, ges < 0.01; spindle density: BF_10_ = 0.34, F_(1)_ = 0.91, p = 0.35, ges = 0.03; slow wave amplitude: BF_10_ = 0.40, F_(1)_ = 0.008, p = 0.93, ges < 0.01; slow wave slope: BF_10_ = 0.35, F_(1)_ = 0.39, p = 0.54, ges < 0.01; slow wave density: BF_10_ = 0.43, F_(1)_ = 0.05, p = 0.83, ges < 0.01; ndPAC: BF_10_ = 0.35, F_(1)_ = 1.34, p = 0.26, ges = 0.05) (see Figure 6). Results on the effects of gender, age and AHI on these metrics are reported in Supplementary Material 6.

**Figure 6.**
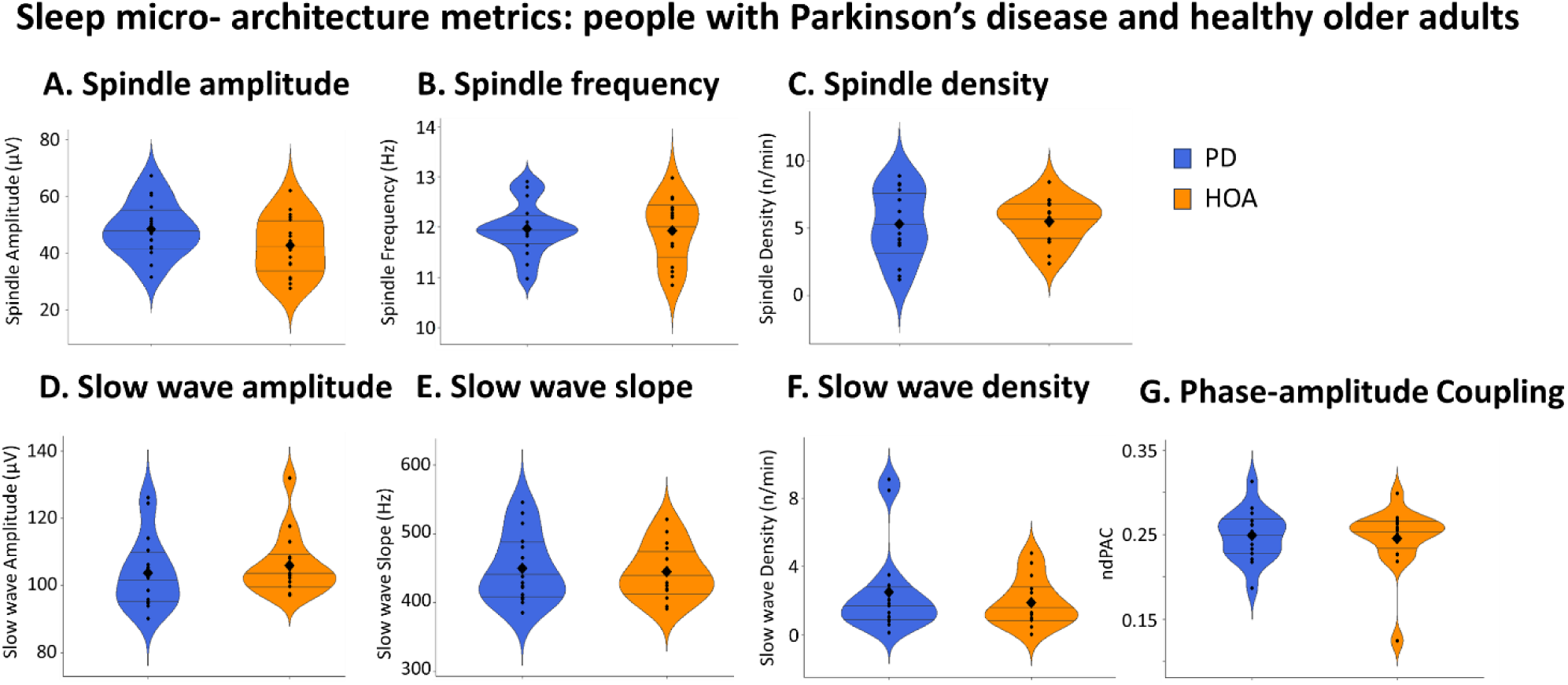
Overview of sleep micro-architecture metrics of people with PD (blue) and HOA (orange) allocated to the nap intervention. (A) Spindle amplitude, (B) spindle frequency, (C) spindle density, (D) slow wave amplitude, (E) slow wave slope and (F) slow wave density; (G) phase-amplitude coupling, measured as normalised direct phase-amplitude coupling (ndPAC). Violin plot: mean (diamond), median (central horizontal bar), and 25^th^ (lower bar) and 75^th^ (higher bar) percentiles.

Lastly, correlations between post-intervention offline changes in PI and sleep micro-architecture metrics obtained from the experimental nap were explored. In PD, moderate evidence for a positive correlation was found between the pre-post-nap relative change in PI and slow wave amplitude (BF_10_ = 5.32, S = 330, r = 0.51, p = 0.04, Figure 7D, in blue), as well as slow wave slope (BF_10_ = 7.08, S = 386, r = 0.43, p = 0.10, Figure 7E, in blue). In contrast, there was weak evidence for no correlation of these metrics in HOA (slow wave amplitude: BF_10_ = 0.63, S = 466, r = 0.17, p = 0.55; slow wave slope: BF_10_ = 0.58, t_(13)_ = -0.50, r = -0.14, p = 0.62, Figure 7D-E, in orange). The comparison of these correlation coefficients revealed no significant difference between groups neither for slow wave amplitude (Z = -0.38, CI [-0.80 – 0.54], p = 0.70), nor for slow wave slope (Z = - 1.02, CI [-1.00 – 0.34], p = 0.31).

**Figure 7.**
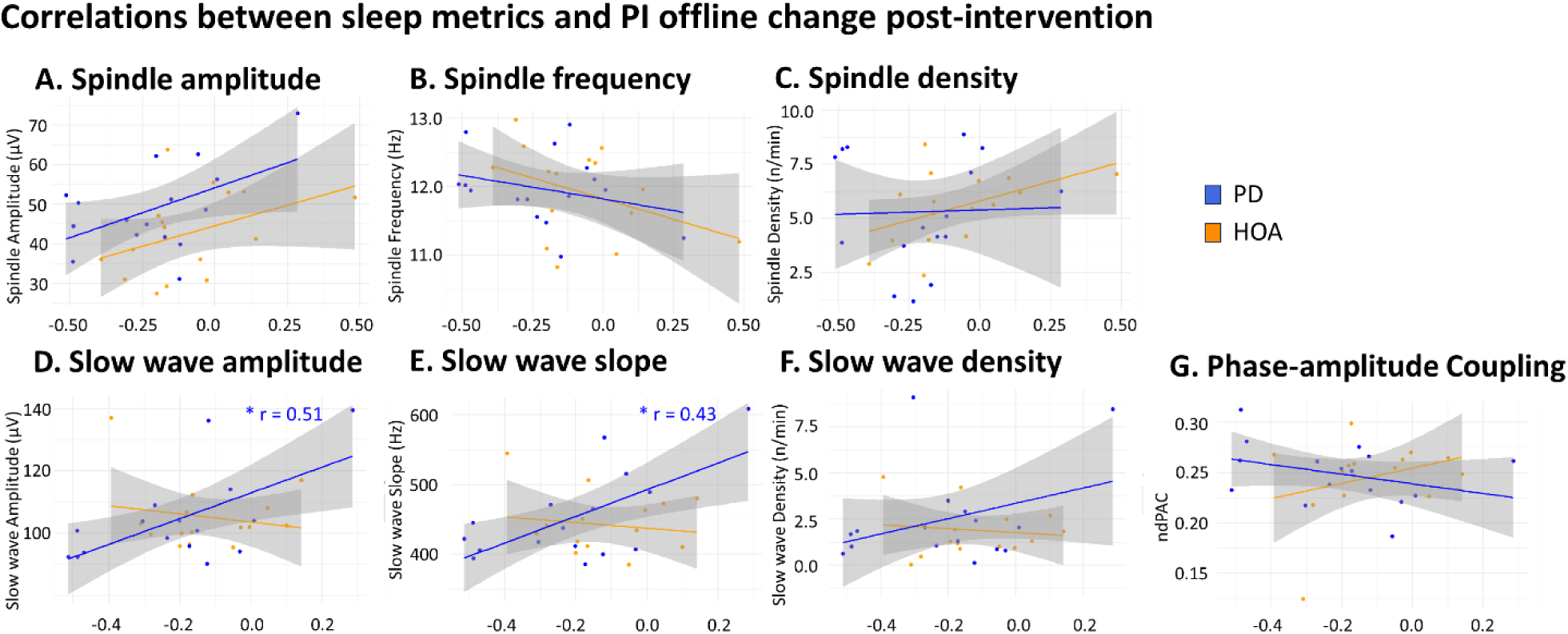
Correlations between sleep micro-architecture metrics and PI offline relative change post-intervention in PD (blue) and HOA (orange) allocated to the nap intervention. (A) Spindle amplitude, (B) spindle frequency, (C) spindle density, (D) slow wave amplitude, (E) slow wave slope and (F) slow wave density; (G) phase-amplitude coupling, measured as normalised direct phase-amplitude coupling (ndPAC). X axis indicating PI offline change post-nap. Dots represent individual values, with linear regression lines for PD (blue) and HOA (orange). The discrepancy between non-significant correlation values and inclined regression slopes may arise due to non-linearity in the data, present specifically in the slow wave density and slow wave amplitude in the HOA group. “*” = moderate evidence for a correlation (3 ≤ BF < 10).

Spindle metrics showed weak evidence in favour of existing correlations in HOA (amplitude: BF_10_ = 1.31, t_(14)_ = 1.68, r = 0.41, p = 0.12; frequency: BF_10_ = 1.27, t_(14)_ = -1.64, r = -0.40, p = 0.12; density: BF_10_ = 1.75, t_(14)_ = 1.95, r = 0.46, p = 0.07, Figure 7A-C, in orange), but in PD this was the case only for spindle amplitude (BF_10_ = 2.30, t_(14)_ = 2.19, r = 0.51, p = 0.05, Figure 7A, in blue), while spindle frequency and density showed weak evidence for no correlation (frequency: BF_10_ = 0.79, t_(14)_ = -1.10, r = -0.28, p = 0.29; density: BF_10_ = 0.52, t_(14)_ = 0.13, r = 0.03, p = 0.90, Figure 7B-C, in blue). Comparisons between groups confirmed no difference between groups’ correlations (spindle amplitude: Z = -0.16, CI [-0.65 – 0.56], p = 0.88; spindle frequency: Z = -0.36, CI [-0.75 – 0.53], p = 0.72; spindle density: Z = 1.19, CI [-0.27 – 1.02], p = 0.24). On the contrary, the amount of N2-N3 sleep showed weak evidence for a correlation with PI changes in PD (BF_10_ = 1.65, t_(14)_ = -1.90, r = -0.45, p = 0.08), but this was weakly favouring no association in HOA (BF_10_ = 0.57, t_(14)_ = 0.43, r = 0.12, p = 0.68). When compared, these correlations evidenced no significant difference between groups (Z = 1.54, CI [-0.16 – 1.13], p = 0.12).

Findings on the relationships between offline changes and slow wave density and ndPAC highlighted weak evidence suggesting no correlation between these metrics, neither in PD (slow wave density: BF_10_ = 0.98, S = 572, r = 0.16, p = 0.56; ndPAC: BF_10_ = 0.99, t_(14)_ = -1.38, r = -0.35, p = 0.19, Figure 7E-F, in blue) nor in HOA (slow wave density: BF_10_ = 0.57, t_(13)_ = -0.44, r = -0.12, p = 0.67; ndPAC: BF_10_ = 0.82, S = 442, r = 0.21, p = 0.45, Figure 7E-F, in orange). Comparisons between these correlation coefficients confirmed no difference between groups (slow wave density: Z = -0.15, CI [-0.76 – 0.65], p = 0.88; ndPAC: Z = 1.37, CI [-0.23 – 1.12], p = 0.17). These results suggest that moderate associations between behavioural performance change on the PI and slow wave amplitude and slope may exist in PD, but we did not find differences with HOA.

## Discussion

In this study, we investigated the effect of a 2-hour post-learning nap compared to wakefulness on the consolidation, 24-hour retention and automaticity of a unimanual self-initiated motor sequence learning task in people with Parkinson’s disease and healthy older adults. Our findings showed that people with PD and HOA exhibit similar levels of consolidation when measured with the performance index. Further, no impact of napping was observed on immediate consolidation, nor on 24-hour retention or automaticity, neither in PD nor in HOA. Although PD had worse subjective sleep, both groups achieved similar sleep efficiency and similar sleep metrics (duration, spindle and slow wave features) were observed during the post-learning nap. Interestingly, positive associations between slow-wave characteristics associated to memory consolidation and performance were observed in the PD group, but they differed from HOA only for the association between phase-amplitude coupling and improvements in accuracy.

The present study indicates that consolidation does not differ between PD and HOA, suggesting that such processes are similarly impaired in both populations [16,87–89]. Further, we found that a 2-hour nap did not improve offline behavioural performance compared to wakefulness, neither in PD nor in HOA. This finding is in agreement with our hypothesis, where we expected similar change in PD and HOA after a diurnal napping intervention. This expectation was based on the partial preservation of sleep-related consolidation in HOA compared to young healthy adults, although not tested in this study [20,21] and the similar findings on sleep-related consolidation in PD and HOA [23–25]. Our results also showed that motor performance did not deteriorate immediately after a period of wakefulness in PD, contradicting our hypothesis that wakefulness would exacerbate the memory consolidation issues, given that the striatum seems particularly implicated in time-facilitated consolidation. This may be due to the fact that, in the current experiment, tests were performed ON dopaminergic medication and the duration of the consolidation period was limited to two hours [1,12].

PD and HOA also performed similarly at post-night retention following the nap/wake intervention, measured with PI. This result is not in line with the findings of Dan and colleagues [25], who compared learning, consolidation and extended practice capacity in *de novo* PD and HOA, finding a worsening of performance in the PD group. Our study adds to the evidence by showing that the implementation of a short sleep intervention after learning does not appear to have an effect on performance after 24 hours in PD ON-medication. However, our findings showed a weak difference in accuracy between PD and HOA at post-night retention in favour of the PD group, suggesting that the latter may have used a different strategy than HOA favouring accuracy over speed in order to achieve similar performance. Preference for accuracy over speed is frequently observed in PD, and likely relates to the core symptom of bradykinesia and to an implicit decision not to move faster because of an already higher than normal energy expenditure to move at a normal speed [90]. Similar results have been reported by Lanir-Azaria and colleagues [24], who investigated whether memory of a practised motor sequence would transfer to another similarly trained sequence, following overnight sleep. Their findings also indicated an improvement in accuracy at post-night retention on all the different initially trained motor sequences, both in PD and HOA.

Delving in the effects of sleep on automaticity, we found that dual-task performance was similar between PD and HOA. This result is not in line with our expectation, given that several studies suggest that automaticity is impaired in PD [91–95]. This could indicate that the secondary counting task used in this study may have not evoked large dual-task costs compared to the single-task alone, allowing people with PD to use compensatory neural networks to achieve apparently similar performance levels on the dual-task [91,96]. Although the possible use of compensation could not be verified in the present study, further practice reduced dual-task costs regardless of the intervention (nap or wake) in both groups, implying that automaticity may not be influenced by post-learning sleep, but rather by online learning [16,17,19,97,98]. Importantly, all participants exhibited initial motor learning. This is in line with current literature suggesting that learning of a motor task is not necessarily impaired in the early to mid-stages of PD [3] possibly due to the activation of compensatory neural circuits [99,100].

Additionally, we found that people with PD and HOA present similar improvement rates with extended practice after a 2-hour nap/wake opportunity, regardless of the intervention. At post-night we found that the people with PD who napped, performed the task more accurately than the HOA, and this group difference was not evident in those who remained awake. Such differential effects of sleep on extended practice in the two groups is in contrast with the findings of Terpening et al. (2013), who demonstrated accuracy improvement rates after a full night of sleep in HOA, but not in PD [23]. In contrast, speed, as well as PI improvement rates of the typed sequence were larger in HOA compared to PD at post-night, regardless of napping. Together, these results suggest that a night of sleep after motor sequence learning may be beneficial to improve speed in HOA, while it may benefit accuracy more in PD.

Interestingly, PD and HOA showed similar sleep quality and quantity of consolidation-related electrophysiological markers during the experimental nap, which is surprising, given the substantial differences on subjective sleep complaints between the groups. It must be noted, however, that in this study we measured a short diurnal sleep period with limited REM sleep, so there was less opportunity for common sleep disorders of PD to impact on sleep efficiency and consolidation [101]. An interesting finding is that performance change after the intervention, as measured with the PI, was associated with higher slow wave amplitude and slope in the PD group only. This is in line with a previous study showing similar correlations in the declarative memory domain in PD [36], but comparisons between correlations from the two populations showed no difference. Moreover, although no association between cross-frequency coupling of spindles and slow waves and better performance was found in either group as measured with PI, HOA did show a positive correlation between cross-frequency coupling and accuracy, in contrast to PD. The same trend was found for speed, although these correlations did not differ statistically between groups (Supplementary Material 4). It thus appears that slow wave amplitude and slope is particularly associated to better performance in PD, whereas cross-frequency coupling seems to benefit performance accuracy in HOA specifically. These findings are in agreement with the evidence suggesting that cross-frequency coupling is important for memory consolidation in the healthy population [29,102], but these speculations may not be extendable to the PD population. Future research is required to determine whether sleep interventions (e.g., targeted memory reactivation) can leverage these specific sleep waves to improve memory consolidation in people with PD. Such non-invasive interventions have already proved their efficacy in young adults [103,104], and are feasible also in older adults and possibly in PD [105–107].

This study presents several limitations. Firstly, despite reaching the anticipated sample of 64 participants (32 people with PD and 32 HOA), consistent with the current literature, the trial was intended as a phase-II explorative study. As such, it may have been underpowered to detect true effects. However, the findings reported can now be used to conduct a sample size calculation for future phase-III studies aiming to investigate the effect of post-learning napping on motor memory consolidation in PD and HOA. Secondly, although we intentionally opted for a napping paradigm to allow for a wake-control group and to minimise possible confounding circadian and homeostatic effects, it is possible that a 2-hour sleep opportunity was too short to show a differential effect compared to wakefulness on MSL performance. Importantly, this study highlights the feasibility of using napping protocols to investigate memory processes in PD, as there was good sleep efficiency and no adverse events recorded.

To conclude, this study showed no beneficial effect of a 2-hour post-learning sleep opportunity as compared to equivalent time of wakefulness on motor memory consolidation in people with Parkinson’s disease and in healthy older adults and no difference between these groups. However, the association between slow wave characteristics and motor behaviour in PD, and not in HOA, may indicate that equivalent performance output could be achieved using different consolidation mechanisms in each group.

## Data Availability

All data produced in the present study are available upon reasonable request to the authors

## Acknowledgements

Funding for this study was provided by the Funds Malou Malou & Perano, managed by the King Baudouin Foundation (2019-J4121350-212854) and Internal Funds KU Leuven (STG/21/035). Moran Gilat was supported by a Marie-Skłodowska-Curie Action Postdoctoral Fellowship (TARGET-SLEEP, 838576). Letizia Micca is supported by a FWO PhD Fellowship (SLEEP ON IT, 11PQB24N). Judith Nicolas was funded by The Fondation Pour la Recherche Médicale (ARF202309017485). The authors would like to sincerely thank all the participants for their efforts. We also like to thank the staff at the Centre for Sleep Monitoring of UZ Leuven, Tine Wouters, Charlotte Moris, Evi Van de Velde and the master thesis students of Rehabilitation Sciences at KU Leuven for their assistance with data collection.

## Data availability statement

The dataset for this study is available upon reasonable request and provision of ethical approval for re-use of the data and data sharing agreement.

