## Supplementary material for "The effect of a post-learning nap on motor memory consolidation in people with Parkinson’s disease: a randomised controlled trial"

#### 1. Methods

##### Motor tasks detailed description

*Psychomotor vigilance test (PVT).* During this 10-minutes sustained attention task, participants were placed in front of a computer screen showing a "+" sign. As soon as the symbol was substituted by a millisecond counter, they had to press on the response button (spacebar), to stop the counter and their reaction time was displayed on the screen. The counter appeared 100 times within 10 minutes, at random intervals between 2 and 10 seconds. In the event that the participants pressed the response button when the counter had not appeared, the "+" sign would automatically decrease its size by 10%, after the message "watch out the counter" appeared at the centre of the screen.

#### 2. Performance Index supplementary results

##### Performance plateau

To test whether performance reached plateau level before the intervention, we explored the differences in performance index (PI) among the four blocks of test pre-intervention with a Bayesian ANOVA, including group (i.e., people with Parkinson disease (PD) and healthy older adults (HOA)), intervention and block as fixed effects and subject as random effect. This was supplemented with a repeated measures ANOVA including the same fixed and random effects. Here we found decisive evidence for an effect of block ($BF_{10} > 1000$, $F_{(3,180)} = 15.62$, $p < 0.01$, p(GG) < 0.01, ges = 0.01), and post-hoc analyses revealed decisive evidence for a difference between the first block and the other three (block 1-2: $BF_{10} = 322.29$, V = 422, $p < 0.01$; block 1-3: $BF_{10} > 1000$, V = 338, $p < 0.01$; block 1-4: $BF_{10} > 1000$, V = 289, $p < 0.01$), but weak to moderate evidence for no difference among the other three (block 2-3: $BF_{10} = 0.86$, V = 839, p = 0.18; block 2-4: $BF_{10} = 0.63$, V = 812, p = 0.13; block 3-4: $BF_{10} = 0.14$, V = 1025, p = 0.92). Weak to moderate evidence was found for no main effects of group and intervention (group: $BF_{10} = 0.98$, $F_{(1,60)} = 2.70$, p = 0.11, ges = 0.04; intervention: $BF_{10} = 0.62$, $F_{(1,60)} = 0.48$, p = 0.49, ges < 0.01), and for their interactions (group by intervention: $BF_{10} = 0.68$, $F_{(1,60)} = 0.26$, p = 0.62, ges < 0.01; group by block: $BF_{10} = 0.14$, $F_{(3,180)} = 1.31$, p = 0.27, p(GG) = 0.27, ges < 0.01; intervention by block: $BF_{10} = 0.16$, $F_{(3,180)} = 1.43$, p = 0.23, p(GG) = 0.24, ges < 0.01; group by intervention by block: $BF_{10} = 0.11$, $F_{(3,180)} = 0.28$, p = 0.84, p(GG) = 0.81, ges < 0.01).

We similarly explored the differences among the first four blocks of training immediately following the intervention, using a Bayesian ANOVA. Findings suggest decisive evidence for an effect of block ($BF_{10} > 1000$, $F_{(3,180)} = 2.39$, $p < 0.01$, p(GG) < 0.01, ges = 0.04), and post-hoc analyses revealed decisive evidence for a difference between the first block and the other three (block 1-2: $BF_{10} > 1000$, V = 199, $p < 0.01$; block 1-3: $BF_{10} > 1000$, V = 245, $p < 0.01$; block 1-4: $BF_{10} > 1000$, V = 240, $p < 0.01$), moderate evidence for a difference between blocks 2 and 4 ($BF_{10} = 3.31$, V = 589, $p < 0.01$) and weak evidence for no difference among the other comparisons (block 2-3: $BF_{10} = 0.38$, V = 872, p = 0.26; block 3-4: $BF_{10} = 0.70$, V = 631, $p < 0.01$). Weak to moderate evidence was found for no main effects of group and intervention (group: $BF_{10} = 0.77$, $F_{(1,60)} = 1.72$, p = 0.19, ges = 0.03; intervention: $BF_{10} = 0.44$, $F_{(1,60)} < 0.01$, p = 0.98, ges < 0.01), and for their interactions (group by intervention: $BF_{10} = 0.56$, $F_{(1,60)} = 0.02$, p = 0.89, ges < 0.01; group by block: $BF_{10} = 0.22$, $F_{(3,180)} = 1.96$, p = 0.12, p(GG) = 0.14, ges < 0.01; intervention by block: $BF_{10} = 0.11$, $F_{(3,180)} = 1.24$, p = 0.30, p(GG) = 0.30, ges < 0.01; group by intervention by block: $BF_{10} = 0.10$, $F_{(3,180)} = 0.22$, p = 0.88, p(GG) = 0.82, ges < 0.01).

On the whole, these findings suggest a first block effect, therefore we decided to exclude this first block of test pre-intervention and the first block of training post-intervention in the calculation of offline performance changes post-intervention, both in PD and in HOA.

#### Supplementary material

The same analysis was performed on the four blocks of test post-intervention, and on the first four blocks or training post-night, used for the calculation of the performance changes post-night, relative to post-intervention performance. On the four blocks of test post-intervention, one subject in the PD group allocated to wake performed no correct sequence in the third block, therefore their data was excluded from this analysis. Here again we found decisive evidence in favour of a main effect of block ($BF_{10} > 1000$, $F_{(3,177)} = 1.67$, $p < 0.01$, p(GG) < 0.01, ges = 0.02). Post-hoc analyses revealed decisive evidence in favour of a difference between the first block and the other three (block 1-2: $BF_{10} > 1000$, V = 339, $p < 0.01$; block 1-3: $BF_{10} = 735.18$, V = 426, $p < 0.01$; block 1-4: $BF_{10} > 1000$, V = 387, $p < 0.01$), and weak to moderate evidence in favour of no difference for the other pairwise comparisons (block 2-3: $BF_{10} = 0.14$, V = 1066, p = 0.69; block 2-4: $BF_{10}$ =0.31, V = 849, p = 0.20; block 3-4: $BF_{10} = 0.39$, V = 836, p = 0.24). For the other factors of the Bayesian ANOVA, weak evidence was found for an effect of group ($BF_{10} = 1.11$, $F_{(1,59)} = 3.10$, p = 0.08, ges = 0.05), while it was weakly to moderately in favour of no effect of intervention ($BF_{10} = 0.56$, $F_{(1,59)} = 0.33$, p = 0.57, ges < 0.01), group by intervention ($BF_{10} = 0.60$, $F_{(1,59)} = 0.005$, p = 0.94, ges < 0.01) group by block ($BF_{10} = 0.11$, $F_{(1,177)} = 0.96$, p = 0.41, p(GG) = 0.40, ges < 0.01), intervention by block ($BF_{10} = 0.22$, $F_{(1,177)} = 1.94$, p = 0.12, p(GG) = 0.14, ges < 0.01), group by intervention by block ($BF_{10} = 0.14$, $F_{(1,177)} = 0.50$, p = 0.63, p(GG) = 0.63, ges < 0.01). The analysis performed on the first four blocks of training post-night revealed decisive evidence in favour of a main effect of block ($BF_{10} > 1000$, $F_{(3,180)} = 30.34$, $p < 0.01$, p(GG) < 0.01, ges = 0.03). Post-hoc analyses revealed decisive evidence in favour of a difference between the first block and the other three (block 1-2: $BF_{10} > 1000$, V = 289, $p < 0.01$; block 1-3: $BF_{10} = 735.18$, V = 200, $p < 0.01$; block 1-4: $BF_{10} > 1000$, V = 204, $p < 0.01$). Block 2 was different from block 3 with strong level of evidence ($BF_{10} = 13.74$, V = 598, $p < 0.01$), but this difference was only weakly evident between block 2 and 4 ($BF_{10}$ =1.78, V = 677, p = 0.02). Blocks 3 and 4 were similar, with supporting moderate level of evidence ($BF_{10} = 0.17$, V = 1086, p = 0.76). Weak evidence was found for an effect of group ($BF_{10} = 1.65$, $F_{(1,60)} = 4.28$, p = 0.04, ges = 0.06), while it was weakly to moderately in favour of no effect of intervention ($BF_{10} = 0.53$, $F_{(1,60)} = 0.21$, p = 0.65, ges < 0.01), group by intervention ($BF_{10} = 0.67$, $F_{(1,60)} = 0.41$, p = 0.52, ges < 0.01) group by block ($BF_{10} = 0.15$, $F_{(1,180)} = 1.67$, p = 0.18, p(GG) = 0.19, ges < 0.01), intervention by block ($BF_{10} = 0.09$, $F_{(1,180)} = 1.00$, p = 0.39, p(GG) = 0.38, ges < 0.01), group by intervention by block ($BF_{10} = 0.27$, $F_{(1,180)} = 1.72$, p = 0.16, p(GG) = 0.18, ges < 0.01). It appears that for the test blocks post-intervention and for the first training block post-night performance was not stable, however, given the degree of evidence, we opted for excluding the first block of the test post-intervention and the first block of training post-night in the computation of post-night offline changes.

To summarise, the last three blocks of test pre-intervention and the first three blocks following the first, of training post-intervention were used to calculate the offline PI changes. Similarly, the post-night relative changes were calculated using the last three blocks of test post-intervention, and the first three blocks, following the first, of training post-night. For consistency, the values of speed and accuracy of the same performance blocks were used to calculate the respective relative changes, post-intervention and post-night.

##### Motor sequence learning and general motor execution

Firstly, to clarify whether any change in performance was related to actual learning and consolidation, or to mere motor execution improvement, we tested for differences in performance between motor sequence learning (MSL) and the motor execution test (MET). We compared the relative changes in PI on the single-task MSL at the start of the first session and the end of the last session, and the relative changes of MET at the start of the first session and that tested at the end of the last session. Notably, one subject in the PD group wake presented missing data for the MET at retention due to technical issues, therefore their data was not included in this model, and in those for sequence duration and accuracy. The Bayesian ANOVA including group, intervention and task (MSL-MET) revealed strong

### Supplementary material

evidence for an effect of task ($BF_{10} = 78.52$, $F_{(1,59)} = 12.13$, $p < 0.01$, ges = 0.08), specifically highlighting larger performance change in the MSL compared to the MET ($BF_{10} = 33.29$, $V = 1803$, $p < 0.01$, $r_{rb} = 0.40$, $p < 0.01$). Instead, weak to moderate evidence was found for no effect of all the other factors (group: $BF_{10} = 0.24$, $F_{(1,59)} = 0.16$, $p = 0.70$, ges < 0.01; intervention: $BF_{10} = 0.27$, $F_{(1,59)} = 0.43$, $p = 0.51$, ges < 0.01; group by intervention interaction: $BF_{10} = 0.46$, $F_{(1,59)} = 1.08$, $p = 0.30$, ges = 0.01; group by task interaction: $BF_{10} = 0.29$, $F_{(1,59)} = 0.38$, $p = 0.54$, ges < 0.01; intervention by task interaction: $BF_{10} = 0.26$, $F_{(1,59)} = 0.17$, $p = 0.68$, ges < 0.01; group by intervention by task: $BF_{10} = 0.43$, $F_{(1,59)} = 0.56$, $p = 0.46$, ges < 0.01). Overall, these results suggest that the performance change was not only related to general motor execution improvement, but to actual learning, irrespective of group and intervention.

#### Supplementary material

##### 3. Sequence Duration analysis

The overall performance of the PD and HOA groups across the three sessions is represented in Figure S1.

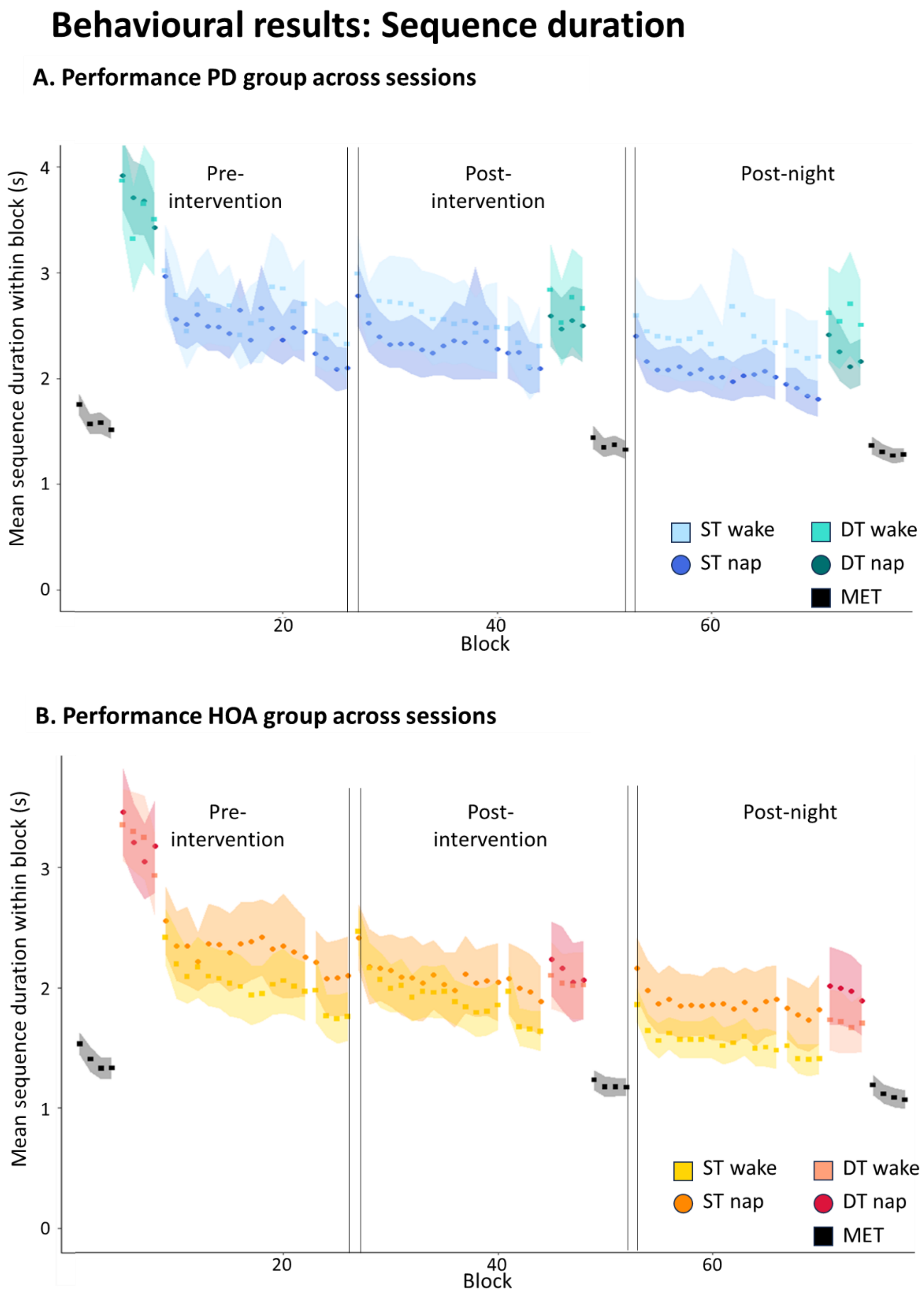


*Figure S1. (A) Representation of sequence duration over the three sessions, expressed as mean and standard error for the PD group (in cold colours) and (B) in the HOA (in warm colours). MET: motor execution test; ST: single task; DT: dual-task.*

### Supplementary material

#### Initial learning

When testing whether the MSL sequence was successfully encoded we found decisive evidence for an effect of block ($BF_{10} > 1000$, $F_{(13,741)} = 7.66$, $p < 0.01$, $p(GG) < 0.01$, ges = 0.01), with an overall trend for improvement during the training, but weak to strong evidence for no effect of all the other main factors and interactions (group: $BF_{10} = 0.69$, $F_{(1,57)} = 0.55$, $p = 0.46$, ges < 0.01; intervention: $BF_{10} = 0.60$, $F_{(1,57)} = 0.02$, $p = 0.88$, ges < 0.01; group by intervention: $BF_{10} = 0.76$, $F_{(1,57)} = 0.69$, $p = 0.41$, ges < 0.01; group by block: $BF_{10} = 0.002$, $F_{(13,741)} = 0.70$, $p = 0.77$, $p(GG) = 0.65$, ges < 0.01; intervention by block: $BF_{10} = 0.007$, $F_{(13,741)} = 0.98$, $p = 0.47$, $p(GG) = 0.44$, ges < 0.01; group by intervention by block: $BF_{10} = 0.28$, $F_{(13,741)} = 1.81$, $p = 0.04$, $p(GG) = 0.10$, ges < 0.01). These results suggest that learning of the MSL occurred over time, but it was not different between populations and intervention groups.

#### Offline changes post-intervention

When comparing offline changes post-intervention, measured with sequence duration, in people with PD allocated to nap and to wake, we found overall weak to moderate evidence for no effect of group, intervention ($BF_{10} = 0.95$, $F_{(1,60)} = 3.21$, $p = 0.08$, ges = 0.05) and their interaction ($BF_{10} = 0.95$, $F_{(1,60)} = 3.21$, $p = 0.07$, ges = 0.05) (Figure S2A).

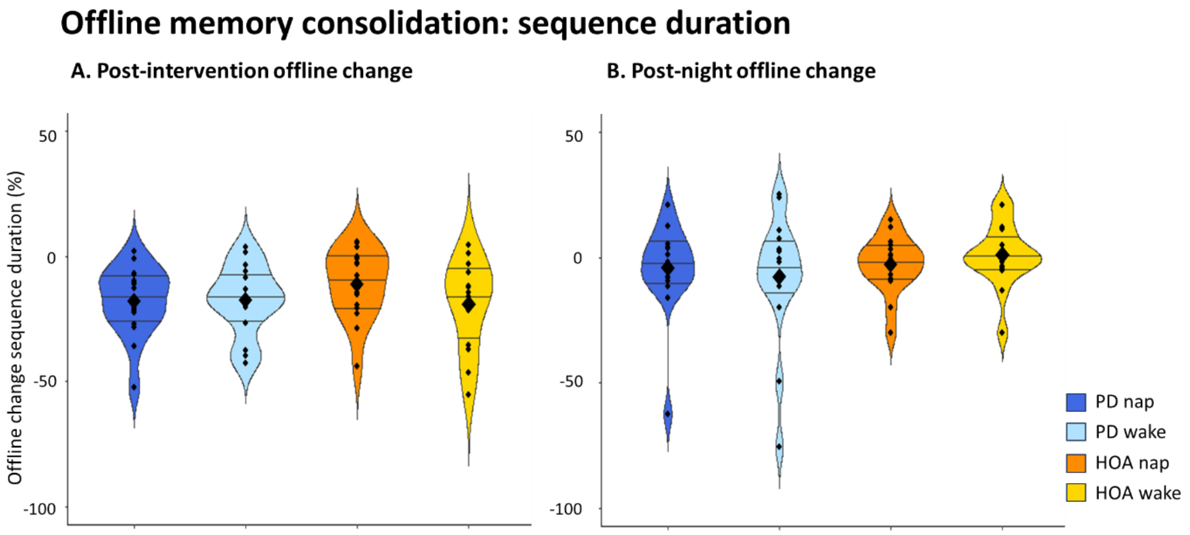


*Figure S2. Offline changes of single-task sequence duration of the MSL sequence in the PD group (blue shades) and HOA (orange shades) (A) post-intervention and (B) post-night.*
*Violin plot: mean (diamond) median (central horizontal bar), and 25th (lower bar) and 75th (higher bar) percentiles.*

#### Post-night offline changes

Our secondary interest was the post-night offline change of the MSL sequence. For this analysis we found weak to moderate evidence for no effect of group ($BF_{10} = 0.45$, $F_{(1,60)} = 1.32$, $p = 0.25$, ges = 0.02), intervention ($BF_{10} = 0.29$, $F_{(1,60)} = 0.29$, $p = 0.59$, ges < 0.01) and of their interaction ($BF_{10} = 0.39$, $F_{(1,60)} = 1.00$, $p = 0.32$, ges = 0.01) (Figure S2B).

#### Dual-task cost

The findings on automaticity measured with sequence duration at post-intervention suggested weak to moderate evidence for no effect of group ($BF_{10} = 0.35$, $F_{(1,60)} = 0.71$, $p = 0.40$, ges = 0.01), intervention ($BF_{10} = 0.32$, $F_{(1,60)} = 0.50$, $p = 0.48$, ges < 0.01), and of their interaction ($BF_{10} = 0.34$, $F_{(1,60)} = 0.70$, $p = 0.41$, ges = 0.01) (Figure S3A). Similar findings were evidenced at post-night (group: $BF_{10} = 0.39$, $F_{(1,60)} = 1.01$, $p = 0.32$, ges = 0.02; intervention: $BF_{10} = 0.27$, $F_{(1,60)} = 0.11$, $p = 0.74$, ges < 0.01; group by intervention: $BF_{10} = 0.33$, $F_{(1,60)} = 0.63$, $p = 0.43$, ges = 0.01).

### Supplementary material

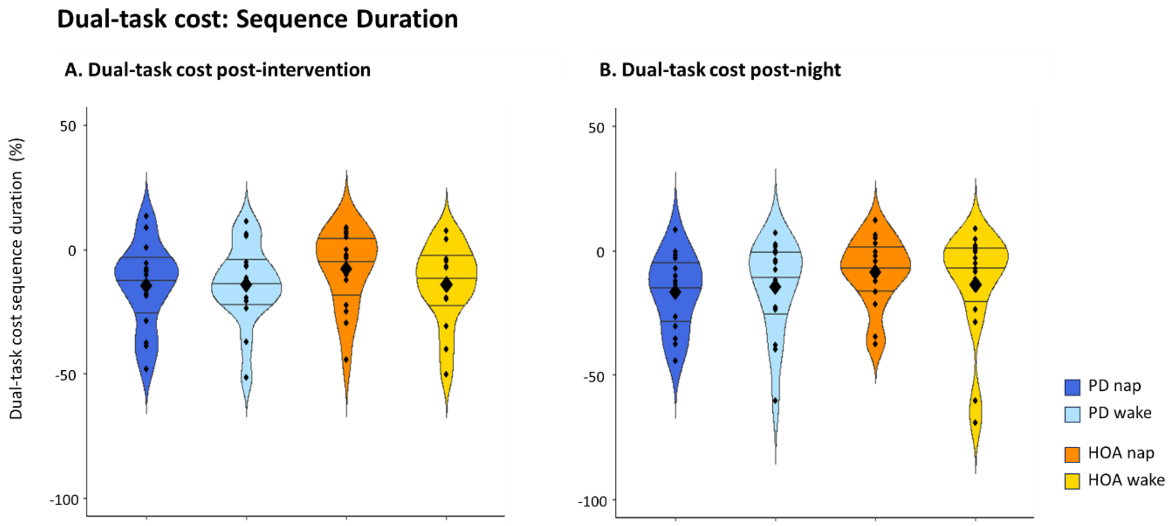


*Figure S3. (A) Post-intervention dual-task cost of sequence duration performance of the MSL sequence in the PD (green shades) and HOA (red shades) groups. (B) Post-night dual-task cost of sequence duration performance of the MSL sequence in the PD and HOA groups*
*Violin plot: mean (diamond), median (central horizontal bar), and 25$^{th}$ (lower bar) and 75$^{th}$ (higher bar) percentiles.*

#### Extended practice post-intervention and post-night

We then tested for the effect of the nap intervention on sequence duration over extended practice post-intervention and post-night. For the analysis we used Bayesian two-way ANOVA's. At post-intervention, we found weak to moderate evidence for no effect of group ($BF_{10}$ = 0.70, $F_{(1,60)}$ = 2.33, p = 0.13, ges = 0.04), intervention ($BF_{10}$ = 0.26, $F_{(1,60)}$ = 0.05, p = 0.82, ges < 0.01) and their interaction ($BF_{10}$ = 0.29, $F_{(1,60)}$ = 0.27, p = 0.61, ges < 0.01). At post-night weak evidence for an effect of group favouring HOA was found ($BF_{10}$ = 1.04, $F_{(1,60)}$ = 3.33, p = 0.07, ges = 0.05), but it was weakly to moderately in favour of no effect of intervention ($BF_{10}$ = 0.26, $F_{(1,60)}$ = 0.004, p = 0.95, ges < 0.01) and their interaction ($BF_{10}$ = 0.45, $F_{(1,60)}$ = 1.39, p = 0.24, ges = 0.02).

In summary, the rate of learning at post-intervention was not different between the populations of study, but it may be larger in HOA at post-night, and it appears that it was not affected by the sleep intervention.

#### Correlation analysis: sequence duration change and sleep micro-architecture

| Correlation parameters | $BF_{10,}$ Correlation test, r, p-value | Correlation comparison |
|---|---|---|
| Sequence duration–NREM2 (%) + NREM3 (%) | PD: $BF_{10}$ = 1.28, $t_{(14)}$ = -1.65, r = -0.40, p = 0.12 | Z = 1.15, CI [-0.30 – 1.03], p = 0.25 |
| | HOA: $BF_{10}$ = 0.53, $t_{(14)}$ = 0.08, r = 0.02, p = 0.94 | |
| Sequence duration–Spindle density | PD: $BF_{10}$ = 0.63, $t_{(14)}$ = -0.73, r = -0.19, p = 0.48 | Z = 1.64, CI [-0.13 – 1.16], p = 0.10 |
| | HOA: $BF_{10}$ = 1.39, $t_{(14)}$ = 1.73, r = 0.42, p = 0.11 | |
| Sequence duration–Spindle amplitude | PD: $BF_{10}$ = 0.61, $t_{(14)}$ = 0.68, r = 0.18, p = 0.51 | Z = 0.33, CI [-0.57 – 0.78], p = 0.74 |
| | HOA: $BF_{10}$ = 0.84, $t_{(14)}$ = 1.18, r = 30, p = 0.26 | |
| Sequence duration–Spindle frequency | PD: $BF_{10}$ = 0.64, $t_{(14)}$ = -0.76, r = -0.20, p = 0.46 | Z = -0.24, CI [-0.75 – 0.60], p = 0.81 |
| | HOA: $BF_{10}$ = 0.81, $t_{(14)}$ = -1.12, r = -0.29, p = 0.28 | |
| Sequence duration–Slow wave density | PD: $BF_{10}$ = 0.79, S = 498, r = 0.27, p = 0.32 | Z = -0.40, CI [-0.83– 0.56], p = 0.69 |
| | HOA: $BF_{10}$ = 0.73, $t_{(13)}$ = -0.95, r = -0.25, p = 0.36 | |
| Sequence duration–Slow wave amplitude | PD: $BF_{10}$ = 1.33, S = 508, r = 0.25, p = 0.34 | Z = -0.57, CI [-0.89 – 0.51], p = 0.57 |
| | HOA: $BF_{10}$ = 1.28, S = 544, r = 0.03, p = 0.92 | |
| Sequence duration–Slow wave slope | PD: $BF_{10}$ = 1.30, S = 556, r = 0.18, p = 0.50 | Z = -0.34, CI [-0.82– 0.59], p = 0.74 |
| | HOA: $BF_{10}$ = 0.71, $t_{(13)}$ = -0.91, r = -0.24, p = 0.38 | |
| Sequence duration– | PD: $BF_{10}$ = 0.52, $t_{(14)}$ = -0.14, r = 0.04, p = 0.89 | |

### Supplementary material

| | | |
|---|---|---|
| ndPAC | HOA: $BF_{10}$ = 0.55, S = 538, r = 0.04, p = 0.89 | Z = 0.94, CI [-0.38 – 0.99], p = 0.35 |

*ndPAC = normalized direct phase-amplitude coupling*
*Offline changes expressed as the negative values of the relative change in sequence duration*

### Supplementary material

#### 4. Accuracy analysis

Figure S4 shows overall performance of the PD and HOA groups across the three sessions.

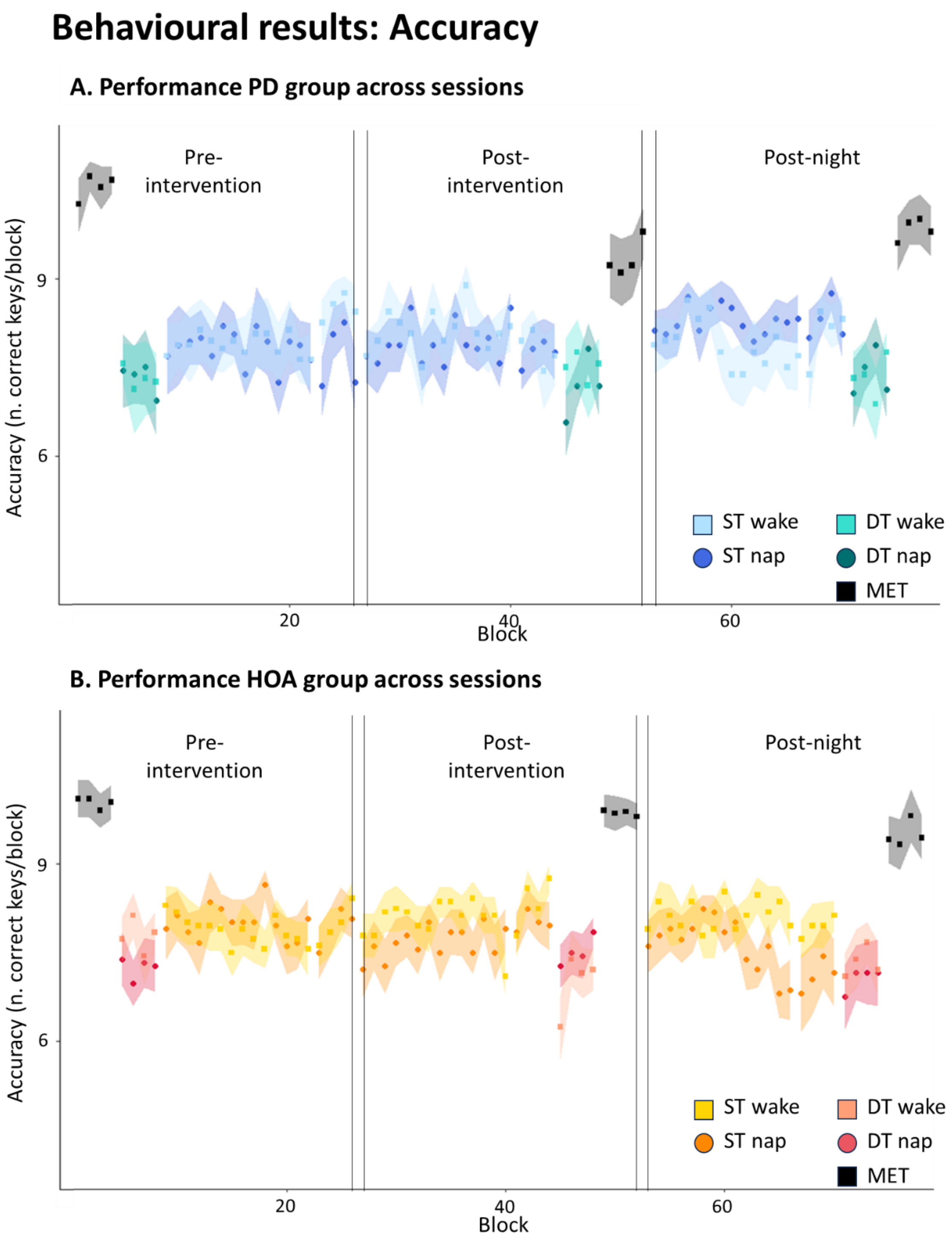


*Figure S4. Representation of performance measured with accuracy over the three sessions, expressed as mean and standard error (A) for the PD group (in cold colours) and (B) for the HOA (in warm colours). MET: motor execution test; ST: single task; DT: dual-task.*

### Supplementary material

#### Initial learning

When testing whether the MSL sequence was successfully encoded we found moderate to strong evidence for no effect of any of the main factors nor their interactions (group: $BF_{10} = 0.25$, $F_{(1,57)} = 0.08$, $p = 0.78$, ges < 0.01; intervention: $BF_{10} = 0.26$, $F_{(1,57)} = 0.19$, $p = 0.66$, ges < 0.01; block: $BF_{10} = 0.002$, $F_{(13,741)} = 1.08$, $p < 0.37$, p(GG) = 0.38, ges < 0.01; group by intervention: $BF_{10} = 0.32$, $F_{(1,57)} = 0.01$, $p = 0.91$, ges < 0.01; group by block: $BF_{10} = 0.01$, $F_{(13,741)} = 1.07$, $p = 0.38$, p(GG) = 0.38, ges < 0.01; intervention by block: $BF_{10} = 0.01$, $F_{(13,741)} = 1.03$, $p = 0.42$, p(GG) = 0.42, ges < 0.01; group by intervention by block: $BF_{10} = 0.02$, $F_{(13,741)} = 0.76$, $p = 0.71$, p(GG) = 0.66, ges < 0.01). These results suggest that while learning of the MSL occurred over time, it was not detectable using accuracy of the typed sequence.

#### Offline changes post-intervention

Weak to moderate evidence for no effect of group ($BF_{10} = 0.28$, $F_{(1,60)} = 0.19$, $p = 0.66$, ges < 0.01), intervention ($BF_{10} = 0.26$, $F_{(1,60)} = 0.07$, $p = 0.79$, ges < 0.01) and of their interaction ($BF_{10} = 0.64$, $F_{(1,60)} = 2.13$, $p = 0.15$, ges = 0.03) was found at post-intervention on the offline changes in accuracy (Figure S5A).

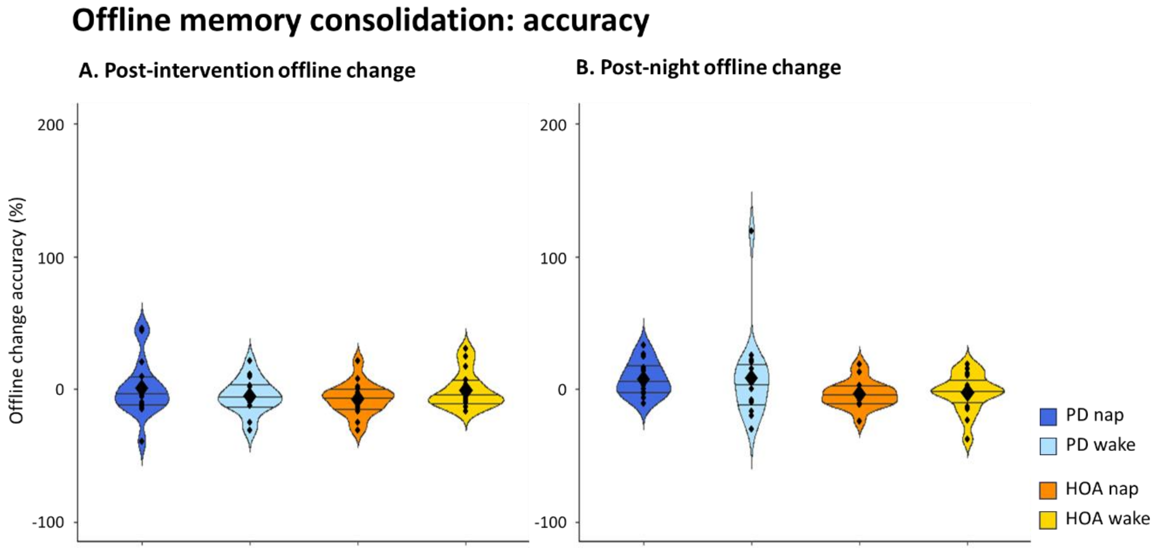


*Figure S5. Offline changes of single-task accuracy of the MSL sequence in the PD group (blue shades) and HOA (orange shades) (A) post-intervention and (B) post-night.*
*Violin plot: mean (diamond) median (central horizontal bar), and 25th (lower bar) and 75th (higher bar) percentiles.*

#### Post-night offline changes

Weak evidence in favour higher offline changes in PD compared to HOA was found at post-night ($BF_{10} = 1.83$, $F_{(1,60)} = 4.62$, $p = 0.04$, ges = 0.07), but it was moderately favouring no effect of intervention ($BF_{10} = 0.28$, $F_{(1,60)} = 0.24$, $p = 0.63$, ges < 0.01) and their interaction ($BF_{10} = 0.30$, $F_{(1,60)} = 0.39$, $p = 0.54$, ges < 0.01) (Figure S5B).

#### Dual-task cost

The findings on automaticity measured with accuracy suggested weak evidence for no effect of group ($BF_{10} = 0.66$, $F_{(1,60)} = 2.35$, $p = 0.13$, ges = 0.04), intervention ($BF_{10} = 0.43$, $F_{(1,60)} = 1.30$, $p = 0.26$, ges = 0.02), and of their interaction ($BF_{10} = 0.87$, $F_{(1,60)} = 3.01$, $p = 0.09$, ges = 0.05) (Figure S6A). At post-night, similar results were found (group: $BF_{10} = 0.73$, $F_{(1,60)} = 2.54$, $p = 0.12$, ges = 0.04; intervention: $BF_{10}$ =

0.40, $F_{(1,60)}$ = 1.11, p = 0.30, ges = 0.02; group by intervention: $BF_{10}$ = 0.65, $F_{(1,60)}$ = 2.29, p = 0.14, ges = 0.04) (Figure S6B).

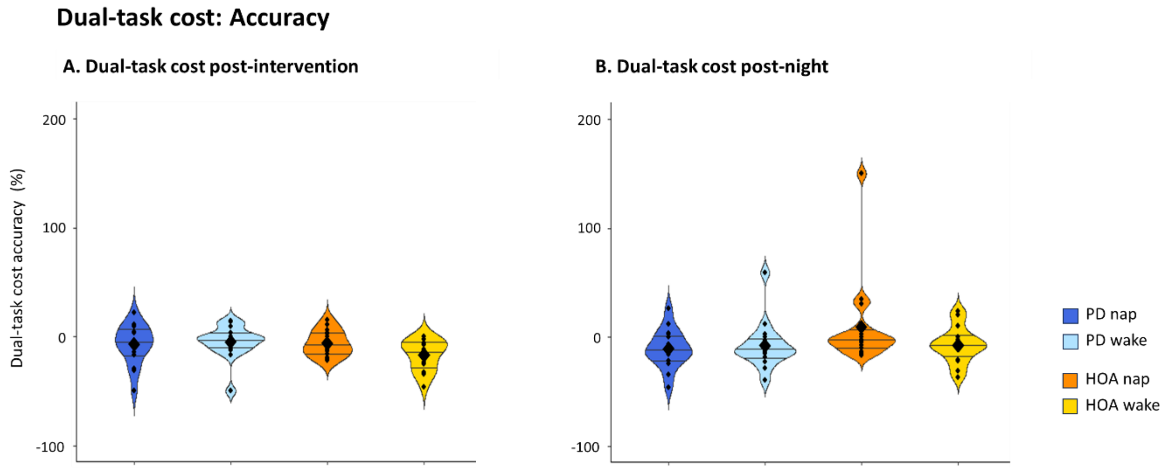


*Figure S6. (A) Post-intervention dual-task cost of accuracy performance of the MSL sequence in the PD and HOA groups. (B) Post-night dual-task cost of accuracy performance of the MSL sequence in the PD and HOA groups.*
*Violin plot: mean (diamond), median (central horizontal bar), and 25th (lower bar) and 75th (higher bar) percentiles.*
*"*" = moderate evidence for a difference (3 ≤ BF < 10); "**" = strong evidence for a difference (10 ≤ BF < 100); "***" = decisive evidence for a difference (> 100)*

#### Extended practice post-intervention and post-night

For this analysis we found weak to moderate evidence for no effect at post-intervention (group: $BF_{10}$ = 0.35, $F_{(1,60)}$ = 0.74, p = 0.39, ges = 0.01; intervention: $BF_{10}$ = 0.28, $F_{(1,60)}$ = 0.23, p = 0.63, ges < 0.01; group by intervention: $BF_{10}$ = 0.26, $F_{(1,60)}$ = 0.07, p = 0.79, ges < 0.01), but at post-night there was moderate evidence in favour of an interaction of group by intervention ($BF_{10}$ = 4.92, $F_{(1,60)}$ = 7.10, p < 0.01, ges = 0.11), while the main effects showed weak to moderate evidence for an effect (group: $BF_{10}$ = 0.27, $F_{(1,60)}$ = 0.13, p = 0.72, ges < 0.01; intervention: $BF_{10}$ = 0.38, $F_{(1,60)}$ = 1.03, p = 0.31, ges = 0.02). Post-hoc tests for the interaction effect revealed weak evidence for a difference between people with PD allocated to the two interventions ($BF_{10}$ = 0.55, W = 149.5, p = 0.43, $r_{rb}$ = -0.15, p = 0.43), while for the HOA we found moderate evidence for a difference favouring the wake intervention ($BF_{10}$ = 4.32, $t_{(30)}$ = -2.66, 95% CI [-2.15– -0.28], p = 0.01, Cohen's d = -0.94). Comparing people with PD and HOA, we found weak evidence for better performance in the people with PD allocated to the nap intervention ($BF_{10}$ = 1.09, $t_{(30)}$ = -1.77, 95% CI [-1.65–0.11], p = 0.09, Cohen's d = -0.63), and moderate evidence for no difference for the participants allocated to the wake intervention ($BF_{10}$ = 0.04, W = 182.5, p = 0.04, $r_{rb}$ = -0.37, p = 0.04).

Hence, accuracy performance change rate during training seemed not to be different between people with PD and HOA, but it appears that a 2-hour nap may have disrupted post-night performance in the HOA.

#### Correlation analysis: accuracy change and sleep micro-architecture

| Correlation parameters | Correlation test, r, p-value | $BF_{10}$ |
|---|---|---|
| Accuracy– NREM2 (%) + NREM3 (%) | PD: $BF_{10}$ = 0.69, $t_{(14)}$ = 0.90, r = 0.23, p = 0.39 | Z = 0.52, CI [-0.48 – 0.81], p = 0.60 |
| | HOA: $BF_{10}$ = 1.27, $t_{(14)}$ = 1.64, r = 0.41, p = 0.12 | |
| Accuracy– Spindle density | PD: $BF_{10}$ = 0.64, $t_{(14)}$ = -0.77, r = -0.20, p = 0.46 | Z = 0.64, CI [-0.48 – 0.90], p = 0.52 |
| | HOA: $BF_{10}$ = 0.53, $t_{(14)}$ = 0.18, r = 0.05, p = 0.86 | |
| Accuracy– Spindle amplitude | PD: $BF_{10}$ = 1.04, $t_{(14)}$ = 1.43, r = 0.36, p = 0.17 | Z = -0.54, CI [-0.84 – 0.49], p = 0.59 |
| | HOA: $BF_{10}$ = 0.59, $t_{(14)}$ = 0.60, r = 0.16, p = 0.56 | |
| Accuracy– Spindle frequency | PD: $BF_{10}$ = 0.65, $t_{(14)}$ = -0.78, r = -0.20, p = 0.45 | Z = -0.24, CI [-0.75 – 0.60], p = 0.81 |
| | HOA: $BF_{10}$ = 0.82, $t_{(14)}$ = -1.14, r = -0.29, p = 0.27 | |

| | | |
|---|---|---|
| Accuracy– Slow wave density | PD: $BF_{10}$ = 27.59, S = 531.89, r = 0.22, p = 0.42<br>HOA: $BF_{10}$ = 0.66, $t_{(14)}$ = 0.78, r = 0.21, p = 0.45 | Z = -0.06, CI [-0.73 – 0.67], p = 0.95 |
| Accuracy– Slow wave amplitude | PD: $BF_{10}$ = 1.64, S = 394.79, r = 0.42, p = 0.11<br>HOA: $BF_{10}$ = 0.57, S = 536.98, r = 0.04, p = 0.88 | Z = -1.01, CI [-1.00 – 0.34], p = 0.31 |
| Accuracy– Slow wave slope | PD: $BF_{10}$ = 1.03, S = 493.86, r = 0.27, p = 0.31<br>HOA: $BF_{10}$ = 0.53, $t_{(14)}$ = -0.11, r = -0.03, p = 0.92 | Z = -0.85, CI [-0.97 – 0.41], p = 0.40 |
| Accuracy– ndPAC | PD: $BF_{10}$ = 2.55, $t_{(14)}$ = -2.28, r = -0.52, p = 0.04<br>HOA: $BF_{10}$ = 2.77, S = 365.83, r = 0.35, p = 0.21 | Z = 2.52, CI [0.20 – 1.38], p = 0.01* |

*ndPAC = normalized direct phase-amplitude coupling*
** significant for frequentist statistics*

#### 5. Correlations of behavioural outcomes with levodopa equivalent daily dose

To test whether levodopa had an influence on the post-intervention and post-night offline changes, and dual-task costs, we performed a correlation analysis between the levodopa equivalent daily dose (LEDD) and each of these behavioural performance measures, using a Bayesian correlation test. For these control analyses we found weak evidence for a correlation between LEDD and performance measured with PI changes post-intervention in the nap group ($BF_{10}$ = 1.38, $t_{(14)}$ = 1.73, r = 0.42, p = 0.11), and post-night in the wake group ($BF_{10}$ = 1.17, $t_{(14)}$ = -1.56, r = -0.39, p = 0.14). Weak evidence for no correlation with LEDD was found for the other metrics (post-intervention wake: $BF_{10}$ = 0.54, $t_{(14)}$ = 0.33, r = 0.09, p = 0.75; post-night nap: $BF_{10}$ = 0.52, S = 517.52, r = 0.24, p = 0.37; post-intervention dual-task cost nap: $BF_{10}$ = 0.61, $t_{(14)}$ = 0.67, r = 0.18, p = 0.51; post-intervention dual-task cost wake: $BF_{10}$ = 0.52, $t_{(14)}$ = 0.16, r = 0.04, p = 0.88; post-night dual-task cost nap: $BF_{10}$ = 0.56, $t_{(14)}$ = 0.43, r = 0.12, p = 0.67; post-night dual-task cost wake: $BF_{10}$ = 0.57, $t_{(14)}$ = -0.51, r = -0.13, p = 0.62).

Similarly, we found weak evidence for no correlation with sequence duration (post-intervention offline changes nap: $BF_{10}$ = 0.65, $t_{(14)}$ = 0.80, r = 0.21, p = 0.44; post-intervention offline changes wake: $BF_{10}$ = 0.56, $t_{(14)}$ = 0.48, r = 0.13, p = 0.64; post-night offline changes nap: $BF_{10}$ = 0.77, S = 539.69, r = 0.21, p = 0.44; post-night offline changes wake: $BF_{10}$ = 0.81, S = 776.14, r = 0.14, p = 0.60; post-intervention dual-task cost nap: $BF_{10}$ = 0.61, $t_{(14)}$ = 0.66, r = 0.17, p = 0.52; post-intervention dual-task cost wake: $BF_{10}$ = 0.61, $t_{(14)}$ = 0.66, r = 0.17, p = 0.52; post-night dual-task cost nap: $BF_{10}$ = 0.58, $t_{(14)}$ = 0.56, r = 0.15, p = 0.58; post-night dual-task cost wake: $BF_{10}$ = 0.57, $t_{(14)}$ = 0.51, r = 0.14, p = 0.62).

For correlations with accuracy we found weak evidence for no correlation with offline changes at any time point (post-intervention offline changes nap: $BF_{10}$ = 0.97, $t_{(14)}$ = 1.35, r = 0.32, p = 0.20; post-intervention offline changes wake: $BF_{10}$ = 0.92, $t_{(14)}$ = 1.29, r = 0.33, p = 0.22; post-night offline changes nap: $BF_{10}$ = 0.58, $t_{(14)}$ = 0.56, r = 0.15, p = 0.58; post-night offline changes wake: $BF_{10}$ = 0.63, S = 752.16, r = -0.11, p = 0.70; post-intervention dual-task cost nap: $BF_{10}$ = 0.64, , $t_{(14)}$ = 0.77, r = 0.20, p = 0.45; post-intervention dual-task cost wake: $BF_{10}$ = 0.60, S = 675.98, r = 0.006, p = 0.98; post-night dual-task cost nap: $BF_{10}$ = 0.53, $t_{(14)}$ = -20.27, r = -0.07, p = 0.79; post-night dual-task cost wake: $BF_{10}$ = 0.73, S = 780.72, r = -0.15, p = 0.58).

Given that the evidence was overall weak, we decided not to account for LEDD in our analyses.

#### 6. Additional findings on electrophysiological markers of plasticity during sleep and demographics

In the absence of a group effect, we next explored the effect of age and gender on sleep micro-architecture. Decisive evidence in favour of an effect of age was found for slow wave amplitude ($BF_{10}$ =

### Supplementary material

964.25, Spearman S = 7220.6, r = -0.47, $p < 0.01$) and for slow wave slope ($BF_{10}$ = 155.71, Spearman S = 7497.1, r = -0.51, $p < 0.01$), suggesting a decrease with age. Strong evidence for an effect of gender was found for spindle amplitude ($BF_{10}$ = 14.76, $t_{(24)}$ = -3.45, 95% CI [-17.60–4.45], $p < 0.01$, Cohen's d = 1.24), with females showing greater spindle amplitude, but weak evidence of an effect of gender was found for slow wave density ($BF_{10}$ = 1.76). No effects of age or gender were found for the phase-amplitude coupling ($BF_{10}$ > 1). Finally, no effect of AHI on sleep micro-architecture metrics was found during the experimental nap (spindle amplitude: $BF_{10}$ = 0.57, spindle frequency: $BF_{10}$ = 0.34, spindle density: $BF_{10}$ = 0.69, slow wave amplitude: $BF_{10}$ = 0.37, slow wave slope: $BF_{10}$ = 0.37, slow wave density: $BF_{10}$ = 0.68, ndPAC: $BF_{10}$ = 0.40).
